# *Tcte1* knockout influence on energy chain transportation, apoptosis and spermatogenesis – implications for male infertility

**DOI:** 10.1101/2022.11.17.22282339

**Authors:** Marta Olszewska, Agnieszka Malcher, Tomasz Stokowy, Nijole Pollock, Andrea J. Berman, Sylwia Budkiewicz, Marzena Kamieniczna, Hanna Jackowiak, Joanna Suszynska-Zajczyk, Piotr Jedrzejczak, Alexander N. Yatsenko, Maciej Kurpisz

## Abstract

**STUDY QUESTION:** Is *Tcte1* mutation causative for male infertility?

**SUMMARY ANSWER:** Collected data underline the complex and devastating effect of the single-gene mutation on testicular molecular network, leading to male reproductive failure.

**WHAT IS KNOWN ALREADY:** Latest data revealed mutations in genes related to axonemal dynein arms as causative for morphology and motility abnormalities in spermatozoa of infertile males, including dysplasia of fibrous sheath (DFS) and multiple morphological abnormalities in the sperm flagella (MMAF). The nexin-dynein regulatory complex (N-DRC) coordinates the dynein arm activity, and is built from DRC1-DRC7 proteins. DRC5 (TCTE1) – one of N-DRC element, has been already reported as a candidate for abnormal sperm flagella beating, however, only in restricted manner with no clear explanation of respective observations.

**STUDY DESIGN, SIZE, DURATION:** Using CRISPR/Cas9 genome editing technique, mouse knockout line of *Tcte1* gene was created on the basis of C57Bl/6J strain. Then, the mouse reproductive potential, semen characteristics, testicular gene expression level, sperm ATP and testis apoptosis level measurements have been performed, followed by visualization of N-DRC proteins in sperm, and protein modeling *in silico*. Also, a pilot genomic sequencing study of samples from human infertile males (n=248) was applied for screening of *TCTE1* variants.

**PARTICIPANTS/MATERIALS, SETTING, METHODS:** To check the reproductive potential of KO mice, adult animals were crossed for delivery of three litters per caged pair, but no longer than for 6 months, in various combinations of zygosity. All experiments were performed for wild type (WT – control group), heterozygous *Tcte1^+/-^*, and homozygous *Tcte1^-/-^* male mice. Gross anatomy was performed on testis and epididymis, followed by semen analysis. Sequencing of RNA (RNAseq; Illumina) has been done for mice testis tissues. STRING interactions have been checked for protein-protein interactions, based on changed expression level of corresponding genes identified in the mouse testis RNAseq experiments. Immunofluorescence *in situ* staining was performed to detect the N-DRC complex proteins: Tcte1 (Drc5), Drc7, Fbxl13 (Drc6), and Eps8l1 (Drc3) in mouse spermatozoa. To determine the ATP amount in spermatozoa, the luminescence level was measured. Also, immunofluorescent *in situ* staining was performed to check the level of apoptosis via caspase 3 visualization on mouse testis samples. DNA from whole blood samples of infertile males (n=137 non-obstructive azoospermia or cryptozoospermia, n=111 samples with spectrum of oligoasthenoteratozoospermia, including n=47 with asthenozoospermia) has been extracted to perform genomic sequencing (WGS, WES or Sanger). Protein prediction modeling of human identified variants and the exon 3 structure deleted in mouse knockout has been also performed.

**MAIN RESULTS AND THE ROLE OF CHANCE:** No progeny at all was found for homozygous males with revealed oligoasthenoteratozoospermia, while heterozygous animals (fertile) manifested oligozoospermia, suggesting haploinsufficiency. RNA-sequencing of the testicular tissue showed the influence of *Tcte1* mutations on the expression pattern of 21 genes responsible for mitochondrial ATP processing, linked with apoptosis, or spermatogenesis. In *Tcte1^-/-^* males the protein revealed only residual amounts in sperm head nucleus, and was not transported to sperm flagella, as other N-DRC components. Decreased ATP level (2.4-fold lower) was found in spermatozoa of homozygous mice, together with disturbed tail:midpiece ratio, leading to abnormal sperm tail beating. Casp3-positive signals (indicating apoptosis) were observed in spermatogonia only, at similar level in all three mouse genotypes. Mutation screening of human infertile males revealed 1 novel and 5 ultrarare heterogeneous variants (predicted as disease causing) in 6.05% of patients studied. Protein prediction modeling of identified variants revealed changes in the protein surface charge potential, leading to disruption in helix flexibility or its dynamics, thus, suggesting the disrupted TCTE1 interaction with its binding partners located within the axoneme.

**What does this mean for patients?:** Abnormal semen parameters (sperm count, motility and morphology) are known as one of the first symptoms that may be related to male fertility problems.

This study aimed to determine the role of *Tcte1* in male infertility using mouse knockout model. Tcte1 protein is one of the structural elements building N-DRC, a complex within an axoneme that is responsible for coordination of the sperm flagella elements activity, strictly related to sperm motility.

We have found that mutations in mouse *Tcte1* knockout model revealed two phenotypes, dependent on zygosity: infertile oligoasthenoteratozoospermic homozygotes, and fertile oligozoospermic heterozygotic males, suggesting haploinsufficiency mechanism. Pilot study on human samples with *TCTE1* variants revealed a wide spectra of semen quality (from non-obstructive azoospermia, via cryptozoospermia, to severe oligoasthenozoospermia).

Thus, *TCTE1* gene is the next one that should be added to the ‘male infertility list’ because of its crucial role in spermatogenesis influencing the variety of testicular molecular networks (incl. energy machinery processing) and proper sperm function.

## INTRODUCTION

Infertility has been defined by World Health Organization as a social disease concerning approximately 10-18% of couples at reproductive age, which are unable to conceive within 1 year of regular unprotected sexual intercourse [WHO, 2020]. Approximately 7% of males and ∼12% of females worldwide reveal reproductive problems, while the male factor is responsible for approximately 20-30% of all infertility cases [Barratt *et al.,* 2017; Vander Borght and Wyns, 2018; Agarwal *et al.,* 2021]. Male infertility is a disease with multifactorial etiology, including: genetic factors, chromosomal aberrations, epigenetic mutations, hormonal abnormalities, infections or reproductive tract abnormalities [Agarwal *et al.,* 2021; Matzuk and Lamb 2008; Zorrilla and Yatsenko 2013; Krausz and Riera-Escamilla 2018]. Genetic causes of male infertility can be detected in ∼10-15% of cases, while in 25-50% of infertile males the etiology remains unexplained [Agarwal *et al.,* 2021; Matzuk and Lamb 2008; Zorrilla and Yatsenko 2013]. The male infertility phenotype mostly results in decreased semen parameters, including: low sperm concentration, abnormal morphology or diminished motility. The actual standards for genetic testing of male infertility rely on chromosomal screening for structural and numerical abnormalities, followed by analyses of deletions in the AZF (azoospermia factor) regions of the Y chromosome [Hwang *et al.,* 2018]. However, it is still not sufficient due to the fact, that approximately 2,000 genes may be related to spermatogenesis, while only ∼10% of them seems to have documented fertility role, so far [Krausz and Riera-Escamilla 2018; Houston *et al.,* 2021] Recent years NGS technology development, including genomic sequencing of DNA (exome or genome; large cohorts of patients), and RNA (RNAseq, scRNAseq on reproductive tissues), followed by animal models of infertility (mostly mouse models; more than 860 genes evaluated, so far [Mouse Genome Informatics, 24 June 2021]) have started the new era of exploration on the massive scale [Houston *et al.,* 2021; Oud *et al.,* 2021; Wyrwoll *et al.,* 2020; Hardy *et al.,* 2021; Xavier *et al.,* 2021].

Beside the genes related to spermatogenesis and low sperm count, also studies of candidates responsible for diminished sperm motility are pending. Thus, the main structural element responsible for motility – the sperm flagella, has to be of high-quality to guarantee sperm motility and then fertilization success. It is known that mutations in genes encoding factors involved in ciliogenesis process, lead to defects in structure and/or functioning of cilia [Girardet *et al.,* 2019; Sironen *et al.,* 2020; Smith *et al.,* 2021; Kumar and Reiter 2021] There is a plenty of data concerning respiratory cilia, however, still little is known about the axonemal dynein arms functioning in sperm tail. Latest data revealed mutations in genes related to axonemal dynein arms as causative for morphology and motility abnormalities in spermatozoa of infertile males [Girardet *et al.,* 2019; Sironen *et al.,* 2020; Aprea *et al.,* 2021; Lorès *et al.,* 2021; Touré *et al.,* 2021; Martinez *et al.,* 2022]. Two definitions were established: (i) for sperm cells with disorganization in axonemal structures, so called dysplasia of fibrous sheath (DFS) [Chemes *et al.,* 1998] and (ii) for multiple morphological abnormalities in the sperm flagella (MMAF), concerning all possible defects of sperm tail, mainly short and thick flagella, and a lack of the central pair (CP) of microtubules or dynein arms [Khelifa *et al.,* 2014]. Spectrum of abnormal sperm tail phenotypes observed allows qualification of a part of disorders into both listed groups (DFS and MMAF) [Sironen *et al.,* 2020; Touré *et al.,* 2021]

The basic structural element of motile cilia and flagella is axoneme built from nine microtubules arranged in doublets positioned radially and one central pair (CP) of singlet microtubules. This central pair is connected to radial spokes (RS) – protein complexes of mechanic regulatory network responsible for regular waveform beating of the cilia [Gui *et al.,* 2019]. RS are linked to inner dynein arms (IDA) localized within each outer dublet. Each outer doublet carries outer dynein arms (ODA) – multi-subunit motor protein complexes, responsible for generation and regulation of sliding between dynein arms leading to ATP-dependent beating [Satir *et al.,* 2014; Wilson *et al.,* 2015]. The 9+2 basic structure is evolutionary conservative [Girardet *et al.,* 2019; Touré *et al*. 2021; Ishikawa 2017]. Doublets are linked with the nexin-dynein regulatory complex (N-DRC), which coordinates the dynein arm activity and stabilizes the attachment of doublet microtubules. The N-DRC is built from DRC1-DRC7 proteins, among which DRC1, DRC2 and DRC4 are responsible for physical linkage between A and B microtubules of doublets [Gui *et al.,* 2019; Morohoshi *et al.,* 2020; Bowar *et al.,* 2018; Wirschell *et al.,* 2013]. In terms of male infertility, the best studied gene among N-DRC is *DRC7* (MIM:618769), which is amenable for proper assembly of the N-DRC structure (in *Drc7^-/-^*male mice the length of the sperm tail was reduced, followed by abnormalities of sperm head morphology) [Morohoshi *et al.,* 2020]. Mouse model of *Drc6* (*Fbxl13*) gene (MIM:609080) revealed no essential role in sperm formation and motility [Morohoshi *et al*., 2020], however, its disruption together with CEP192 (MIM:616426) leads to defects in HEK293T cell motility [Fung *et al.,* 2018]. Mutations in *Drc3* gene (MIM:618758) indirectly revealed male infertility as a lack of pregnancy success with wild-type females [Ha *et al.,* 2016].

One of the N-DRC structural genes is *TCTE1* (T-Complex-associated-Testis-Expressed 1), known also as *DRC5, FAP155*, *D6S46* in humans (*locus* 6p21.1; MIM:186975) or *Tcte1*, *D17S*, *Tcte-1*, *D17Sil1* in mouse (*locus* 17 B3; 17 22.54 cM; MGI:98640). *TCTE1* is evolutionary conserved in most eukaryotic organisms, that contain motility cilia or flagella in their life cycle (Figure 1), and is required for proper flagella functioning. Expression of *TCTE1* is being observed in testis (highest; with expression starting at the stage of spermatid), choroid plexus and Fallopian tube. TCTE1 contains five leucine rich repeats (LRRs). To date, only two reports described a role of *Tcte1* gene: as a mice model of infertility, where the *Tcte1*-*null* sperm cells exhibited asthenozoospermia or in group of patients without clear explanation of the linkage between observed variant and sperm phenotype [Zhou *et al.,* 2022; Castaneda *et al.,* 2017]. Thus, understanding of the nature of this specific motility type defect in *Tcte1-null* observations has been limited, so far.

**Figure 1:**
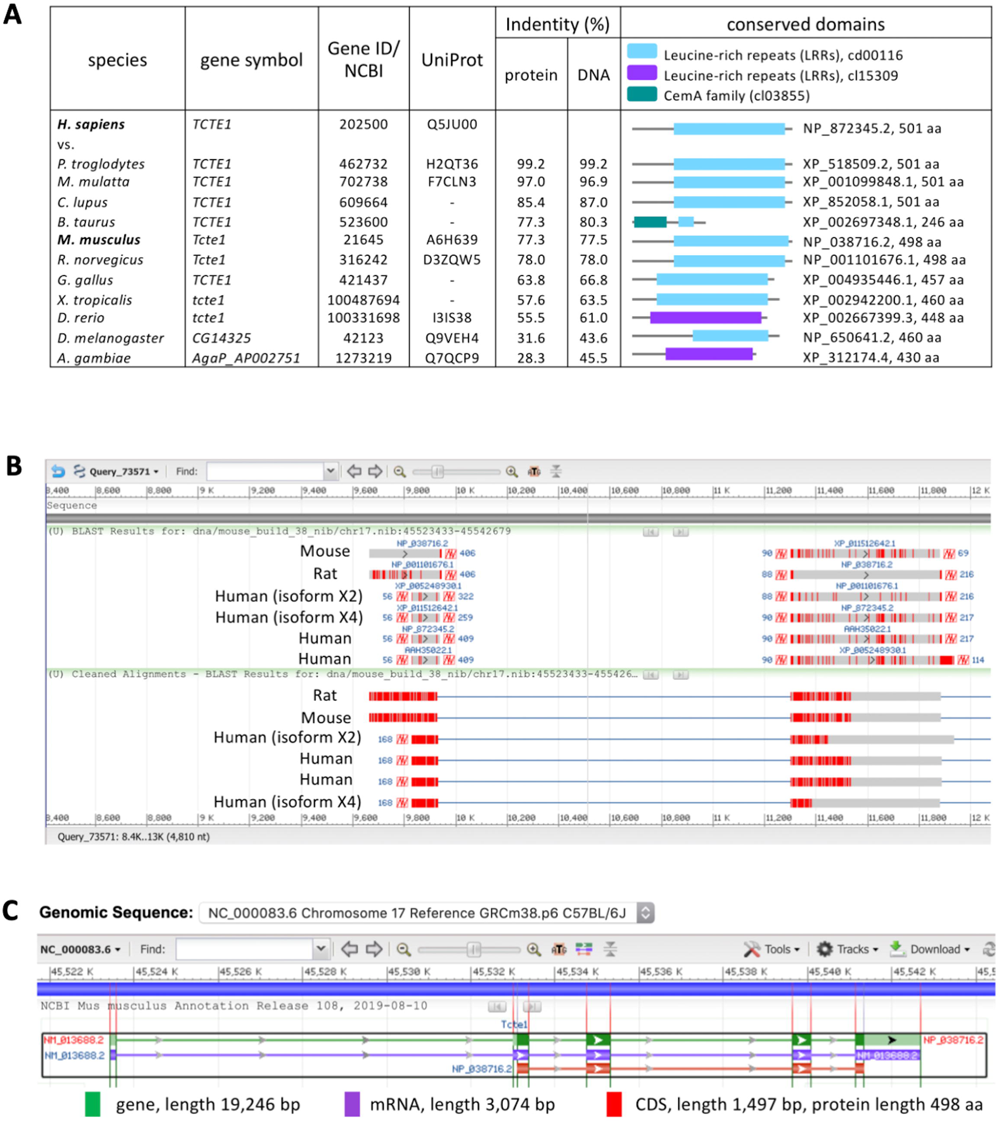
Characteristics of *Tcte1* gene. **A** Putative homologs of *Tcte1* gene conserved in Bilateria, according to HomoloGene (8434), including collation of proteins and their conserved domains; **B** Sequence alignement between human, mouse and rat; **C** NCBI Gene schematic representation of *Tcte1* gene in *Mus musculus*.

In our study we aimed to determine the role of *Tcte1* in male infertility using mouse knockout model. We created homo-and heterozygous animals, which were next examined for their reproductive potential, differences in anatomy and histology, and semen parameters. We also performed RNA sequencing (RNAseq) in testis tissue to obtain an answer about the influence of *Tcte1* null mutation on the expression pattern of other gonadal genes. Also, the ATP measurement of spermatozoa has been performed. Additionally, screening of variants in human samples from males with severely decreased sperm count was made, followed by protein prediction modeling of revealed variants. Thus, this study contains novel and the most complete data concerning *TCTE1* role in male infertility, so far.

## MATERIALS AND METHODS

### Animals

Using CRISPR/Cas9 genome editing technique, mouse heterozygous knockout line of *Tcte1* gene (*Tcte1^+/^*) was created on the basis of C57Bl/6J strain in the Cyagen Biosciences company (USA). The mouse *Tcte1* gene is located on chromosome 17 B3; 17 22.54 cM (GenBank: NM_013688.2; Ensembl: ENSMUSG00000023949). Exon 3 of the *Tcte1* gene was selected as target site (deletion of 1292 bp) (Figure 2A). gRNAs targeting vectors (constructed, sequenced and generated by *in vitro* transcription) were injected into fertilized oocytes (Supplementary file 1). The founders were genotyped and those with positive results were bred to the next generation. Then, in the animal facility of the Institute of Human Genetics PAS (Poznan, Poland), adult *Tcte1^+/-^* animals were mated to generate homozygous *Tcte1^-/-^* mice. Wild type *Tcte1^+/+^*(WT) animals C57Bl/6J were purchased from Charles River company (Germany). Relative expression of *Tcte1* gene was verified using Real-time PCR method (Figure 2B; Supplementary file 2). All mice were housed in pathogen-free animal facility with food and water *ad libitum*, stable temperature of 22°C, and a light:dark light cycle of 12:12. All procedures were in accordance with the guidelines and regulations for animal experimentations and restricted usage of genetically modified organisms (GMO), and were approved by the Local Ethical Committee for Animal Experiments at the Poznan University of Life Sciences (no. 1/2014, 18/2017), and Ministry of Environment (no. 70/2017). Mouse models with infertility phenotypes were reviewed in the Mouse Genome Informatics database (Jackson Laboratory; http://www.informatics.jax.org/) and through literature searches (PubMed; https://pubmed.ncbi.nlm.nih.gov/).

**Figure 2:**
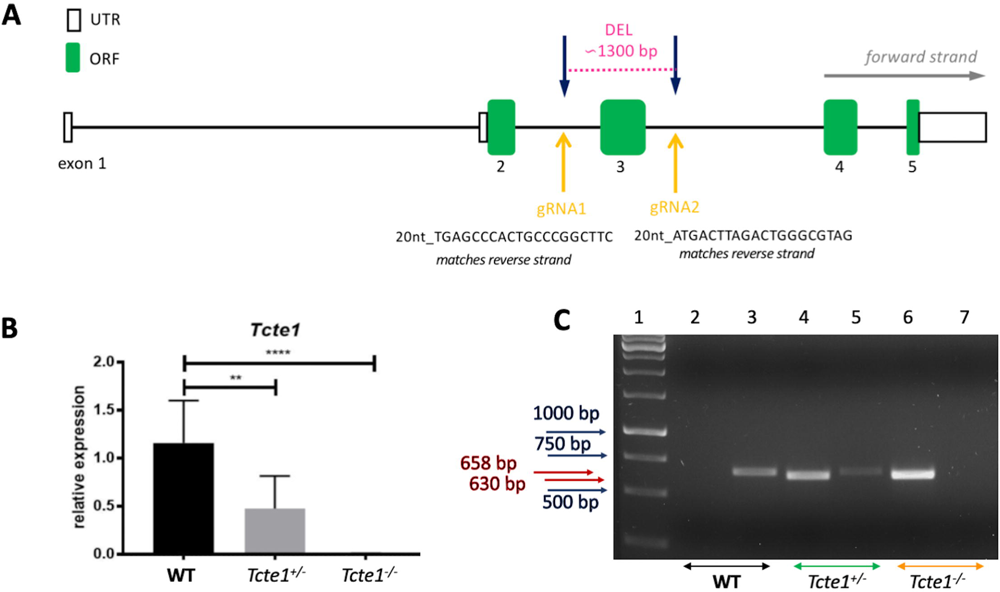
Characteristics of *Tcte1* mice knockout. **A** Schematic representation of gRNA localization in CRISPR/Cas9 KO creation; **B** Relative expression of *Tcte1* gene in wild type (WT), hetero- ^(+/-)^ and homozygous mice ^(-/-)^ (Real-time PCR); **** p<0.0001, ** p<0.01; **C** Example of genotyping result. Lane 1 – 1kb marker (GeneRuler), lanes 2, 3 – wild type (WT), lanes 4, 5 – hetero- ^(+/-)^, and lanes 6, 7 – homozygous mice ^(-/-)^; lanes: 2, 4, 6 identified the lack (0 bp) or presence of mutation (630 bp), lanes: 3, 5, 7 differentiated hetero-(658 bp) and homozygotes (0 bp).

### Reproductive potential

To check the reproductive potential of KO mice, adult animals (7-8 weeks old) were crossed for delivery of three litters per caged pair, but no longer than for 6 months. Crossing combinations included: (i) 4 pairs: homozygous *Tcte1^-/-^* male × wild type (WT) female, (ii) 5 pairs: heterozygous *Tcte1^+/-^* male × heterozygous *Tcte1^+/-^* female, and (iii) 3 pairs: heterozygous *Tcte1^+/-^* male × wild type (WT) female. Vaginal plug presence was checked every morning to confirm mating incidence. Time period from mating start point to first pedigree appearance was checked. Number of pups was counted on the day after birth. For each crossed pair, the number of litters, pups, and sex ratio were estimated (also according to pups’ genotypes) (Supplementary file 3).

### Genotyping

Genotyping of the pups delivered was performed via PCR assay, with two sets of primers: (i) to identify wildtype and positive KO (het-and homozygous) animals: Tcte1-F: CGTTTTAAGAATGTATTGAGGGTTGGG, Tcte1-R: CTCCGTAGGCTCCTGCCAATATG; estimated product size: for WT 1915 bp, for KO ∼630 bp (deleted ∼1300 bp); PCR results analysis: one band with 1915 bp or 0 bp determined WT, while one band with 630 bp evidenced hetero-or homozygotes; (ii) to distinguish between hetero-and homozygotes: Tcte1-WT/He-F: AGCTTGCCACACCCTCAAGGTACTA, Tcte1-R: CTCCGTAGGCTCCTGCCAATATG; estimated product size: 658 bp for heterozygotes and 0 bp for homozygotes; PCR results analysis: one band with 658 bp determined heterozygotes, while no one band (0 bp) indicated homozygotes (Figure 2C). The PCR mixture and reaction conditions are described in Supplementary file 4.

### Tissue gonadal preparation/collection

Gross anatomy was performed on testis and epididymis of WT (n=21), *Tcte1^+/-^* (n=22), and *Tcte1^-/-^* (n=18) male animals (10 weeks old). Mice were weighted, then euthanized with carbon dioxide (CO_2_) overdose (euthanasia Minerve Easy-box; Minerve; France) with stable flow rate per minute. Testes and epididymides were collected, weighted, and measured. For histological evaluation, two fixing approaches have been applied. Both approaches started with anaesthetization under isoflurane (4%) and transcardial perfusion with 50 ml/animal of ice-cold PBS, followed by 50 ml/animal of 10% formalin solution, neutrally buffered (Sigma Aldrich, USA, cat. no. HT501850) for 3 males per zygosity and technical variant. Next, the tissues have been immersed in formalin solution for 24 hours, 4°C, in darkness. Then, in the first approach, Paraplast-embedded sections were prepared (5 µm thickness), using Microtome Leica 2055. In the second technique, gonads were sequentially immersed in 15%, 20% and 30% of sucrose (Sigma Aldrich, cat. no. S0389), each for 24 hours, at room temperature. Thereafter, gonads were coated with Tissue-Tek O.C.T. Compound (Sakura, cat. no. 4583) and frozen in cold (-80° C) 2-Methylbutane (Sigma Aldrich, USA, cat. no. M32631). Then, cryo-sections were prepared (10 µm thickness), using Leica Cryostat CM1950 and kept at −80°C until further stainings. To obtain the histological picture, slides were stained with Masson-Goldner protocol [Romeis *et al.,* 2010] for classic paraffin blocks or standard progressive H&E staining method for frozen samples. Briefly, after fixation twice in 100% EtOH (10 min.), slides were first hydrated in the EtOH series (95%, 70% for 5 min.), washed in distilled water and stained in Mayer’s hematoxylin (Sigma) for 5 min., then washed in distilled water (3 times, 5 min.) and dehydrated in 50%, 70 %, 80%, 96% EtOH (5 min. each). Counterstaining with eosin Y (Sigma) for 30 seconds was applied, followed by washing in 100% EtOH, clearing in xylene (5 min. each, two repeats), and mounting in DPX (Sigma). Thereafter, slides were documented with Leica DM5500 light microscope (×40 dry or ×63 oil immersive objectives, motorized stage), LASX software (with Navigator tool). All the evaluations have been performed blindly by two experienced scientists per technique.

For each zygosity the geometrical features of the tubule sections have been evaluated, incl.: tubule diameter (D) and cell layer thickness (CL) (the mean of two measurements made at opposite sides of the tubule) and lumen diameter (l), using Fiji (Image J) measurement tool. To check density of the cells, ratios of: D:CL, D:l and CL:l were also estimated. In total, for each zygosity 133 -144 tubule sections were evaluated (3 random sections per zygosity, each min. 30 tubules).

### Mouse semen samples

Mouse spermatozoa were collected from epididymis *post mortem*. Collected epididymis were placed immediately in a 2.0 mL test tube with warm 500 µL of a non-capacitating Whitten’s HEPES medium (pH 7.2-7.4, 37°C), then cutted into 5-7 fragments, and incubated in 37°C for 15 min. to allow the sperm cells to swim out. Whittens-HEPES medium contained: 100.0 mM NaCl, 4.4 mM KCl, 1.2 mM KH2PO4, 1.2 mM MgSO4, 5.4 mM glucose, 0.8 mM pyruvic acid, 4.8 mM lactic acid (hemi-Ca), HEPES 20 mM [Wertheimer *et al.,* 2013]. Then, spermatozoa were collected and proceeded with seminological analysis (concentration, motility, morphology). Next, semen samples were fixed in a fresh fixative solution (methanol:acetic acid, 3:1 v/v, −20°C; 3×) and stored until further use in −20°C.

### Semen analysis

10 μl of sperm suspension was applied on the Makler counting chamber (Sefi medical instrument Ltd., Israel) to test the sperm concentration and total sperm count. Also, the ratio of progressive (fast and slow), non-progressive (total and circular) and immotile spermatozoa were calculated [WHO, 2020]. For assessment of sperm morphology, the Papanicolaou staining was performed. Briefly, the air-dried smears were fixed in ethanol:diethyl ether (1:1 ratio, vol/vol) solution for 10 min. and left for air-drying. Then, slides were stained by Papanicolau procedure according to WHO guidelines [WHO, 2020], and mounted in DPX Mountant (Sigma-Aldrich; USA; cat. no. 06522). A total of 200 spermatozoa were analyzed using Leica DM5500 light microscope (×1000 magnification, oil immersion). Various morphological forms of spermatozoa were determined including abnormalities of head, midpiece and tail. Also, the length of sperm tail, midpiece and the tail:midpiece ratio were measured to check possible differences between males with various zygosity (6 males/per zygosity); for each male at least 100 spermatozoa were measured with the ‘length measurement’ option by CellSense Dimension software (Olympus).

### Sequencing of RNA (RNAseq)

For RNA extraction testis tissue was collected into in RNAlater buffer (Invitrogen; Waltham, MA, USA) and freezed immediately (-80°C). Samples were processed using the AllPrep DNA/RNA/Protein Mini Kit (Qiagen; Hilden, Germany). Quality and quantity of extracted RNA samples were checked using NanoDrop (Thermo Scientific) and Quantus (Promega), and 1 μg of RNA per sample has been used for sequencing (Macrogen; South Korea). RNA sequencing was performed with Illumina NovaSeq 6000 platform using TruSeq Stranded mRNA LT Sample Prep Kit, with specified conditions of paired-end reads at 100 bp, and throughput 100 M reads per sample. RNA-Seq raw reads were aligned to mouse reference genome mm10 using hisat 2.0.5 with Gencode vM13 transcriptome reference. Reads aligned in the coding regions of the genome were counted using Feature Counts. Then, read counts were normalized using DESeq2 and subjected to differential analysis. The differential analysis included computing of median based fold change, statistical significance using the Students’ t-test and false discovery rates (correction for multiple testing) in the R/Bioconductor programming environment. Testis tissue expression was retrieved from GTEx (https://gtexportal.org/home/), BioGPS (http://biogps.org/), and AceView (https://www.ncbi.nlm.nih.gov/IEB/Research/Acembly/). All RNA-seq expression data have been deposited in Gene Expression Omnibus (GEO), accession number GSE207805.

### STRING interactions

For analysis of protein-protein interactions, based on changed expression level of corresponding genes identified in the mouse testis RNAseq experiments, the STRING analysis was performed [version 11.0]. STRING database allows to delineate probability of the interactions between evaluated proteins using different evidence channels [Mering *et al.,* 2005; Szklarczyk *et al.,* 2019]. This tool enables the integration of all data available in public resources concerning protein-protein interactions.

### Immunofluorescence on sperm – N-DRC visualization

Immunofluorescence *in situ* staining was performed to detect the N-DRC complex proteins: Tcte1 (Drc5), Drc7, Fbxl13 (Drc6), and Eps8l1 (Drc3) in mouse spermatozoa. Mouse spermatozoa were collected and fixed as described above. Specific antibodies were applied: primary antibodies – rabbit anti-human-TCTE1 (cat. no. orb357083), rabbit anti-mouse-Drc7 (orb586958), anti-mouse-Fbxl13 (orb314611), and anti-mouse-Eps8l1 (orb382538; all from Biorbyt; Cambridge, UK); secondary antibody – goat anti-rabbit-AF594 (Abcam; cat. no. ab150160; Cambridge, UK). The UniProt IDs and homology of the immunization regions of primary antibodies between human and mouse were described in Supplementary file 5. Antibodies were diluted in 0.5% PBST (primary: 1:100, secondary: 1:500). First, slides with fixed sperm smears were washed in a series of washes in 1× PBST, and then incubated in 25 mM DTT/1 M Tris-HCl, pH 9.5, at room temperature for 20 min. Then, washing in 1× PBST, and incubation in 6 N HCl for 30 min. was applied, followed by blocking step with 1% BSA/1× PBST for 30 min. Next, overnight incubation with antibodies was performed at 4°C in a humidified container. After washing, the samples with 1× PBST, secondary AF594-conjugated antibody was applied for 1 hr. Next, unconjugated antibodies were washed out (4× in 1× PBST, 5 min each, slight rotation). For the final detection, 20 ul of Fluoroshield with DAPI (Sigma-Aldrich; cat. no. F6057) was added to the samples, and microscopic analysis was performed. Images were acquired using a fluorescence microscope with a proper filter set: Leica DM5500, filters: DAPI/TxR/Triple; objectives: 10× and 63× with oil immersion; DFC 7000T camera; software: LASX.

### Sperm ATP measurement

For measurement of ATP in spermatozoa the CellTiter-Glo 2.0 Assay (Promega; USA) was used. This ready-to-use assay allows to determine the quantitation of the ATP amount, indicating the presence of metabolically active cells. The assay is based on the thermostability of luciferase (Ultra-GloTM Recombinant Luciferase), and was performed as described in manufacturers’ protocol. Briefly, all laboratory work was done in the stable temperature of 22°C. After collection of mouse spermatozoa (as described above), the amount of approximately 1× 10^6^ of sperm cells was transferred to black opaque 1.5 mL test tube and proper amount of CellTiter-Glo kit was added (1:1, v:v). Next, 2 min. of orbital mixing of the content was performed to activate cell lysis. Then, after 10 min. of incubation (to stabilize the luminescence), the measurement of luminescent signal was completed using Glomax 20/20 luminometer (Promega; USA). For each sample, three measurements (repeats) were done, each in two time points of 0 and +60 min. Numbers of animals used were: WT (n=7), *Tcte1^+/-^* (n=7), and *Tcte1^-/-^* (n=6). Mean luminescence values obtained (represented as RLU – relative luminescence unit) were compared between tested mice groups.

### Immunofluorescence on testis

Immunofluorescence *in situ* staining on testis was performed to check: (i) mean cell count of spermatogonia and spermatocytes (preleptotene and pachytene) in tubules’ sections (cryo-sections), and (ii) to estimate the level of apoptosis via caspase 3 visualization on mouse testicular samples (paraffine embedded sections) from all the three zygosities. Caspases play important role in the induction, transduction, and amplification of intracellular apoptotic signals, and thus, caspase-3 can be used as one of the apoptotic indicators [Lej et al, 2022]. Mouse testes were collected and fixed as described above. Specific antibodies were applied: for (i) primary antibodies – mouse anti-MLH1 (Abcam; Cambridge, UK, cat. no. ab14206), 1:30 in 0.5% PBST, rabbit anti-RAD51 (Abcam; Cambridge, UK, cat. no. ab63801), 1:200 in 0.5% PBST; (ii) primary antibody – rabbit anti-Casp3, (Biorbyt; Cambridge, UK, cat. no. orb10237), 1:100 in 0.5% PBST; secondary antibodies – goat anti-mouse-FITC (Sigma-Aldrich, cat. no. F2012), 1:400 and goat anti-rabbit-AF594 (Abcam; Cambridge, UK, cat. no. 150080), 1:500 in 0.5% PBST. First, slides with paraffine fixed testis cross sections were deparaffinized in 3 changes of xylene (10 mins. each), while for cryo-sections this step has been omitted. Then slides were incubated in a series of ethanol: 100%, 96%, and 70%, twice for 5 mins. in each solution. Next, after rinsing in distilled water, tissue sections were incubated sequentially in: a freshly prepared 2% NaBH_4_/1× PBS, and in 0.1M glycine (30 min., room temp. each). Heat-induced epitope retrieval was performed by immersing the slides in a 0.01M sodium citrate/0.05% Tween 20, pH 6.0 for 30 min., followed by cooling down in a room temp. for next 20 min. Blocking step was done in an aliquot of freshly prepared 1% BSA/1× PBST, at 37 °C for 1 h. Then, tissue sections were incubated with primary antibody in a humidified container overnight at 4°C. Secondary antibody was applied for 1 hr, followed by the washing out of the unconjugated antibody (4× in 1× PBST, 5 min each, slight rotation). For the final detection, 20 μl of Fluoroshield with DAPI (Sigma-Aldrich; cat. no. F6057) was added. Images were captured using a fluorescence Leica DM5500 microscope with a proper filter set: DAPI/TxR/SpG/Triple; objectives: 10× and 63× with oil immersion; DFC 7000T camera, LASX software. The analysis was performed for 3 sections per zygosity, each min. 30 tubules (for cell count) or 2 cross-sections per zygosity, 15-20 tubules per tissue section (for Casp-3 evaluation).

### TUNEL assay

TUNEL assay was performed to check the level of apoptosis and necrosis on the testicular tissue according to the manufacturer’s protocol (Abcam, ab66110 TUNEL Assay Kit – BrdU-Red). Breifly, paraffine embedded sections (or 2 cross-sections per zygosity, with 15-20 random tubules analyzed/slide) have been deparaffinized in xylene (twice, each 5 min.), followed by EtOH series: 100% (twice, 5 min.), 95%, 85%, 70% and 50% (3 min.). Next, slides were incubated in 0.85% NaCl (5 min.) and washed in PBS. After incubation with Proteinase K solution (20 ug/mL in Tris-HCl-EDTA; 5 min.) and washing with PBS, slides were immersed in 4% formaldehyde/PBS. Then, Wash Buffer was applied twice for 10 min. with plastic coverslip on the top. Next, DNA Labeling Solution was added and incubation in a dark humidified incubator for 1,5 hr in 37°C was processed. After next two washes with PBS, Antibody Solution was applied and slides were incubated for 30 min. at room temp. Last steps included: ddH_2_O washing (5 min., twice) and application of 20 μl of Fluoroshield with DAPI (Sigma-Aldrich; cat. no. F6057) for the final detection. Slides were analyzed within 1 hr using the fluorescent Leica DM5500 microscope with a proper filter set: DAPI/TxR/Triple; objectives: 10× and 63× with oil immersion; DFC 7000T camera, LASX software.

### Sequencing of the human samples

A pilot genomic sequencing study of samples from human infertile males was also applied. Bioethical committee approvals were received for the study (Local Bioethics Committee of Poznan University of Medical Sciences, approval no. 1003/18; National Bioethical Committee, Ministry of Health, Warsaw, Poland, approval no. OKB-5-2/15). All participants were notified about the aim of the study, and provided written informed consent. All experiments were performed in accordance with relevant guidelines and regulations. DNA from whole blood samples (collected into sterile single test tubes with EDTA) was extracted using standard protocols of MagCore HF16 Automated Nucleic Acid Extractor (RBC Biosciences, Taiwan), or Gentra Puregene kit (Qiagen, USA). Total number of sequenced patients was n=248, including: 137 with non-obstructive azoospermia (lack of spermatozoa in ejaculate) or cryptozoospermia (spermatozoa detectable in ejaculate after centrifugation), followed by 111 samples with spectrum of seminal abnormalities in concentration, and/or motility, and/or morphology. Among those samples, 47 revealed decreased motility (asthenozoospermia). Semen evaluation was done according to WHO guidelines [WHO, 2020], followed by AUA and ASRM guidelines for infertile male diagnosis [Schleger *et al.,* 2021]. All men revealed normal hormonal levels, had no chromosomal abnormalities and a lack of AZF microdeletions. For whole exome samples (n=76 from Pittsburgh), sequencing libraries were constructed using the Agilent technologies (Santa Clara, USA), including: SureSelect XT All Exon V4 + UTRs capture kit and SureSelect XT Target Enrichment System. Libraries were sequenced with average read coverage of 62-100x (∼6Gb per sample), using 100 bp paired-end sequencing with TruSeq PE Cluster Kit v3 cBot HS and 4 × 50 TruSeq SBS V3 HS sequencing kit (Illumina, San Diego, USA) on the HiSeq2000 (Illumina, San Diego, CA). Whole genome sequencing (n=42, Poznan) was performed using Illumina HiSeqX with the coverage of at least 30x (100-120 Gb per sample). Sanger sequencing for all exomes of *TCTE1* gene (n=130, Poznan) was performed using LightRun96 system of Eurofins Genomics (Eurofins Genomics AT GmbH, Vienna, Austria). Samples were prepared with QIAquick Gel Extraction (cat. no. 28704) or PCR Purification (cat. no. 28104) kits (Qiagen, USA), and analyzed with CLC Main Workbench 8 software (Qiagen, USA).

### Filtering of whole genome sequencing results

The quality of raw NGS data was evaluated using the FastQC and MultiQC packages. Reads were aligned to the reference human genome GRCh37 using bwa-mem aligner included in the Speedseq package [Chiang *et al.,* 2015]. For calling of single nucleotide variants (SNVs) and small indels the freeBayes v0.9.21 (speedseq-var) was implemented, following previously described pipeline [Supernat *et al.,* 2018]. Variant disease association and missense mutation prediction effects were determined using: dbSNP (https://www.ncbi.nlm.nih.gov/snp/), OMIM (https://www.ncbi.nlm.nih.gov/omim/), ClinVar (https://www.ncbi.nlm.nih.gov/clinvar/), MutationTaster (http://www.mutationtaster.org/), SIFT (https://sift.bii.a-star.edu.sg/), Polyphen (http://genetics.bwh.harvard.edu/pph2/), CADD (https://cadd.gs.washington.edu/snv), MetaDome (https://stuart.radboudumc.nl/metadome/), REVEL (rare exome variant ensemble learner) [Ioannidis *et al.,* 2016], Ensembl Variant Effect Predictor [Mclaren *et al.,* 2016], and Human Gene Mutation Database (HGMD; Qiagen, Redwood City, CA). SNV pathogenicity was assessed on the basis of American College of Medical Genetics and Genomics (ACMG) guidelines [Richards *et al.,* 2015]. PhyloP (Cold Spring Harbor Laboratory) and Clustal Omega were used for determination of conservation (https://www.ebi.ac.uk/Tools/msa/clustalo/). All NGS variants were confirmed via the Integrative Genomics Viewer (Broad Institute). Variant frequencies have been obtained from the gnomAD database (frequencies from more than 125k exomes and 15k genomes, http://gnomad.broadinstitute.org; v2.1.1), and were filtered in search of rare variants (with minor allele frequency (MAF) <1%).

### Protein prediction

The amino acid sequence of TCTE1 was input into Phyre2 [Kelley *et al.,* 2015] for both intensive and fast mode homology modeling, I-TASSER [Roy *et al.,* 2010; Yang *et al., 2*015, Yang and Zhang 2015], and AlphaFold [Jumper *et al.,* 2021; Varadi *et al.,* 2022] which utilize sequence alignment-based algorithms for generating predicted secondary and tertiary structures; AlphaFold additionally uses machine learning to integrate biological principles of protein folding with the alignment-based algorithms. Confidence scores were obtained from these programs; AlphaFold additionally generates predicted aligned error plots for assessing the quality of the model’s long-range interactions. Models were inspected manually and aligned using the secondary structure matching (SSM) [Krissinel and Henrick, 2004] tool in COOT [Emsley *et al.,* 2010] to minimize user input bias. Docking of the structures into electron tomography data was performed manually in COOT [Emsley *et al.,* 2010]. No refinement was performed to minimize bias. All structure model figures and mutations without post refinement, were generated using Pymol (Schrödinger). I-TASSER was used also for prediction of the exon 3 structure deleted in our mouse KO model (Supplementary file 6).

### Statistical analysis

Statistical analyses included the following: normality test (D’Agostino and Pearson or Kolmogorov-Smirnoff), unpaired two-tailed t-test with Welch’s correction or Mann-Whitney test, one-way ANOVA test and Fisher’s exact, and two-tailed Pearson correlation. All tests were performed with a significance level of α=0.05 using GraphPad Prism (v.7.0e) software.

## RESULTS

### Reproductive potential of KO mice

Reproductive potential results were shown in Figure 3 and Supplementary file 3. Only pairs with homozygous *Tcte1^-/-^* males revealed no progeny (*Tcte1^-/-^* male x WT female). Other combinations (*Tcte1^+/-^* male × *Tcte1^+/-^* female; *Tcte1^+/-^* male × WT female) showed similar results, when considering: number of litters, pups, and sex ratio. Mean time of caging to delivery of the 1^st^ pedigree was longer in *Tcte1^+/-^* male × *Tcte1^+/-^* female combination by 25.45% (statistically not significant). When considering genotypes of the pups, statistically significant differences (p<0.05) were observed in the number of *Tcte1^-/-^*male and female pups delivered – no homozygous animals were obtained from combination of *Tcte1^+/-^* male × WT female, while in mating of *Tcte1^+/-^* male × *Tcte1^+/-^* female, homozygous males (33.25% of all pups) and females (24.67%) were received.

**Figure 3:**
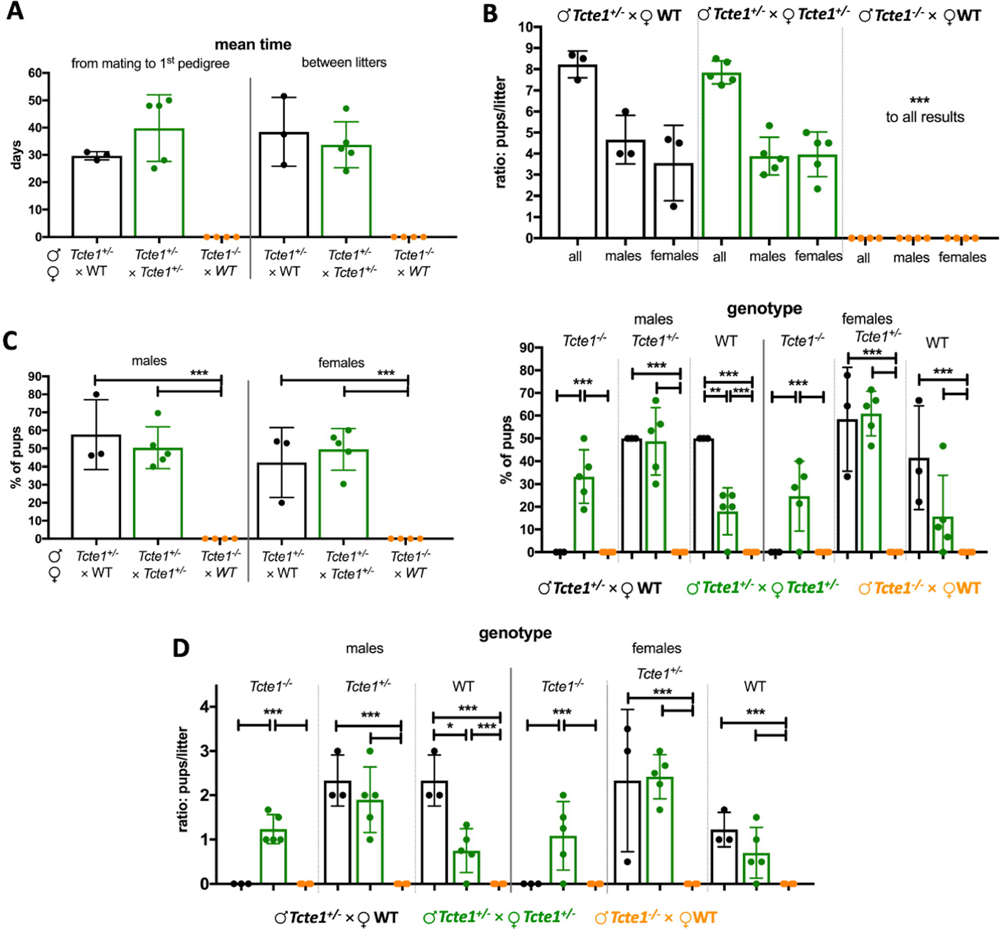
Reproductive potential of KO *Tcte1* mice. Mating combinations and their results including: time of breeding, number of litters and pups, number of male and female pups, and genotypes observed in pups of each combination. **A** Mean time: from mating to 1st pedigree, and between particular litters; **B** Ratio of males and females per litter in each mating combination; **C** Frequency of males and females and their genotype observed each mating combination; left panel: according to sex of pups; right panel: according to mating combinations; **D** Ratio of males and females per litter and their genotype observed each mating combination. *** p < 0.0001, ** 0.0001 < p < 0.01, * 0.01 < p < 0.05

### Histopathology and seminal parameters

Histopathological evaluation of testicular and epididymal sections revealed no visual (structural) differences between WT, *Tcte1^+/-^*, and *Tcte1^-/-^* males confirming preserved spermatogenesis (Figure 4A, Supplementary file 7). No odds were found for body mass of the animals and the length of the epididymis (Figure 4B). In the *Tcte1^-/-^* males testis weight was decreased by approximately 41% (p<0.0001) followed by the ∼22% decline of its size (p<0.0001), while in *Tcte1^+/-^* males only testis weight was slightly decreased (approx. 7%; p<0.05) when compared to control animals. Differences in dimensions of the tubule sections were noted (Figure 4C), with increasing tubule and lumen diameters while decreasing cell layer thickness in *Tcte1^-/-^*. In *Tcte^+/-^* the cell layer thickness and tubule diameter were also increased. When comparing germ cell count (Figure 5) reduced numbers of spermatogonia (∼22%) and pachytene spermatocytes (13-17%) were documented for *Tcte1^-/-^*and *Tcte^+/-^* (p<0.001), while preleptotene spermatocytes level was elevated (p<0.001). Mean sum of evaluated the cell types was similar between WT and homozygotes (65.95 and 64.39 of cells/tubule section, respectively). Such data suggest some kind of time shift at the first stages of prophase – germ cells from KO mice seem to be delayed at preleptotene stage, while WT males enters pachytene stadium at higher ratio.

**Figure 4:**
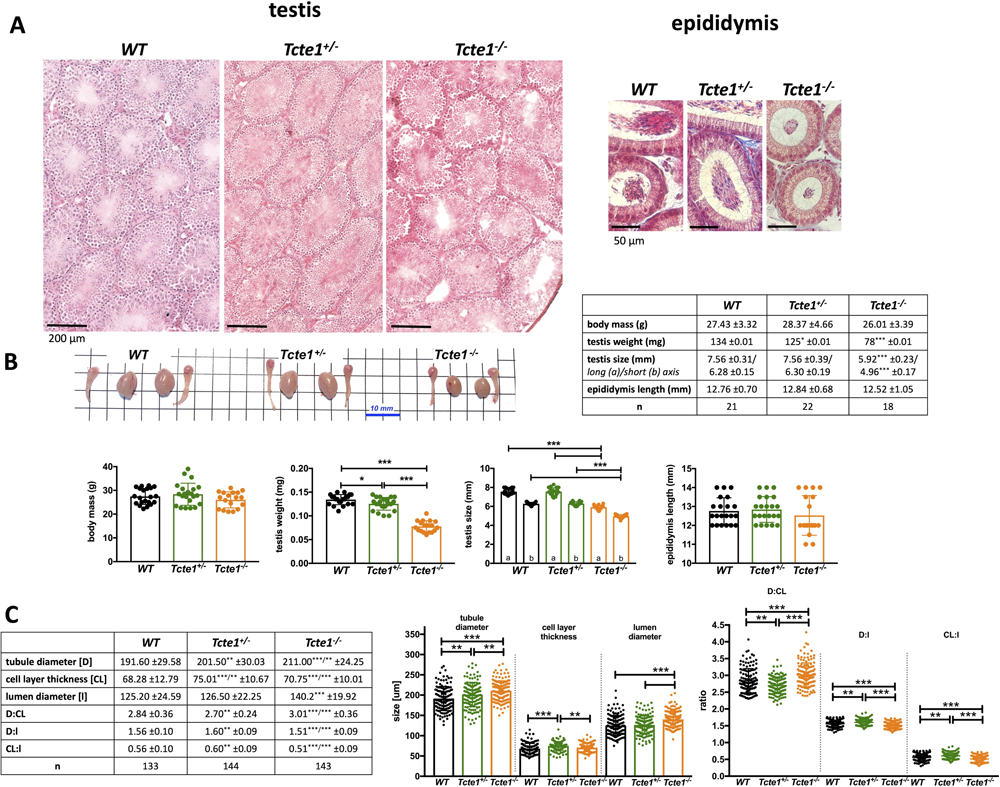
Characteristics of *Tcte1* wild type (WT), hetero- ^(+/-)^ and homozygous mice ^(-/-)^. **A** Histopathology of testes and epididymis (caput); staining: classic hematoxyline-eosine staining (testis) or Masson-Goldner protocol (epididymis); microscope: Leica DM5500, objective 40×, LASX software with Navigator function; **B** Comparison of body mass, testis weight and size, and epididymis length, followed by gross morphology of gonads (n – no. of mice analyzed); **C** Comparison of the dimensions of testicular tubules’ sections (n – no. of sections analyzed); *** p < 0.0001, ** 0.0001 < p < 0.01, * 0.01 < p < 0.05

**Figure 5:**
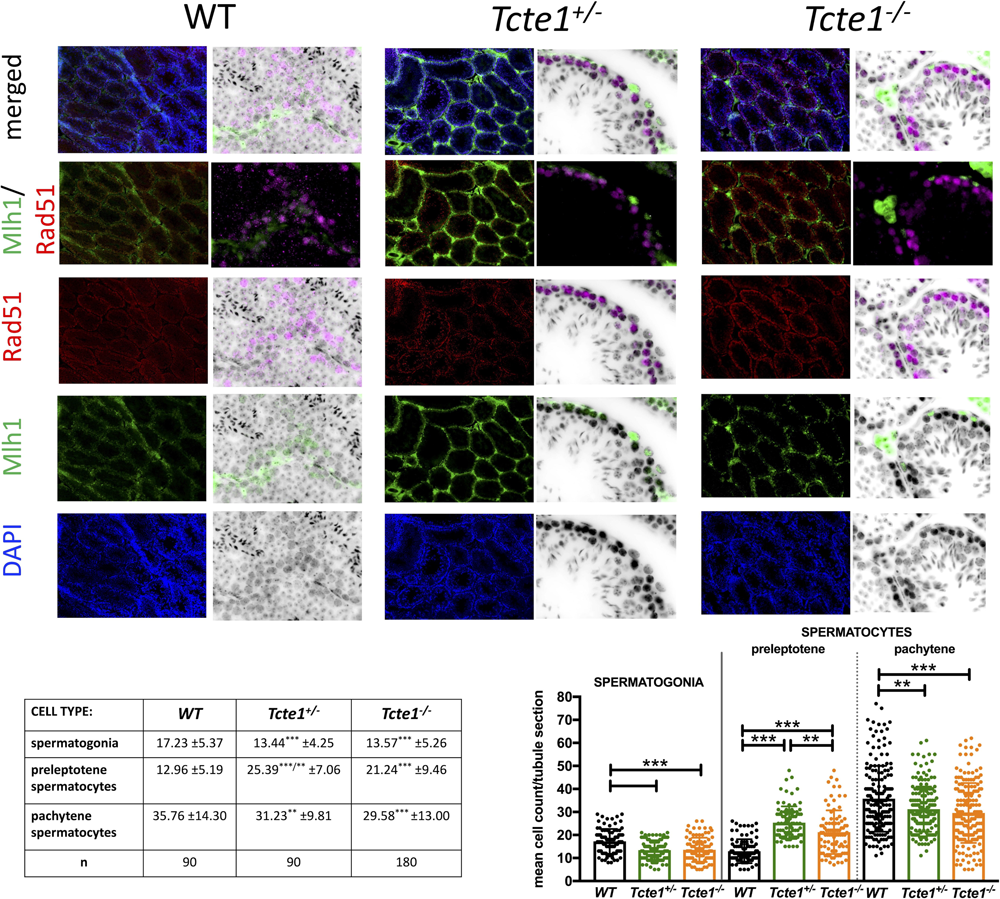
Evaluation of germ cell types in testes of *Tcte1* wild type (WT), hetero- ^(+/-)^ and homozygous mice ^(-/-)^. Three germ cell types were evaluated: spermatogonia (stained with intensive green color), preleptotene spermatocytes (intensive red color), and pachytene spermatocytes (medium green and red). Immunofluorescent staining with antibodies – primary: mouse anti-MLH1 (Abcam; Cambridge, UK, cat. no. ab14206), 1:30, rabbit anti-RAD51 (Abcam; Cambridge, UK, cat. no. ab63801), 1:200; secondary: goat anti-mouse-FITC (Sigma-Aldrich, cat. no. F2012), 1:400 and goat anti-rabbit-AF594 (Abcam; Cambridge, UK, cat. no. 150080), 1:500.

When considering seminal parameters, both: in *Tcte1^-/-^*as well as in *Tcte1^+/-^* males the sperm concentration was importantly decreased (3.27, and 2.47-fold, respectively), when collating to control results (p<0.0001; Figure 6A). Additionally, in *Tcte1^-/-^* males the frequency of progressively motile spermatozoa was residual, with progressive type of motility in only 0.22% of spermatozoa (decreased ∼150-fold vs. control 33.52%, (p<0.0001)), followed by 4.89% of slow progressive spermatozoa (decreased ∼3-fold vs. control 15.50%) (p<0.0001). Interestingly, ∼2.4-fold increase in circular type of motility (around own axis and non-progressive) was observed for *Tcte1^-/-^* males when compared to WT and *Tcte1^+/-^* animals (p<0.01) (Supplementary file 8; Supplementary file 9; Supplementary file 10). The frequency of immotile spermatozoa also increased in *Tcte1^-/-^*males (2-fold to control; p<0.0001). The frequency of morphologically normal spermatozoa in *Tcte1^-/-^* animals was significantly decreased (2-fold to control; p<0.0001; Figure 6A, B, C); Supplementary file 11). The majority of morphological defects was observed for sperm head, mostly of decapitated spermatozoa in *Tcte1^-/-^* animals – 52.55% vs. 19.16% for *Tcte1^+/-^*and 23.33% for WT, and amorphous head shape with decreased frequency in *Tcte1^-/-^*(26.11%) and *Tcte1^+/-^* (33.72%) males vs. WT animals (48.88%) (Supplementary file 11). Midpiece defects were found in *Tcte1^+/-^* (9.71%) and WT (8.20%) animals, while *Tcte1^-/-^* mice revealed only low frequency (0.38%) of such defects. On the other hand, an increased frequency (∼5-fold) of coiled tail was observed for *Tcte1^+/-^* animals when compared to other mice zygosity (p<0.01) (Supplementary file 11). Summarizing, seminological changes in mice with *Tcte1* mutation resulted in oligoasthenoteratozoospermia in homozygous *Tcte1^-/-^* or oligozoospermia in heterozygous *Tcte1^+/-^* animals.

**Figure 6:**
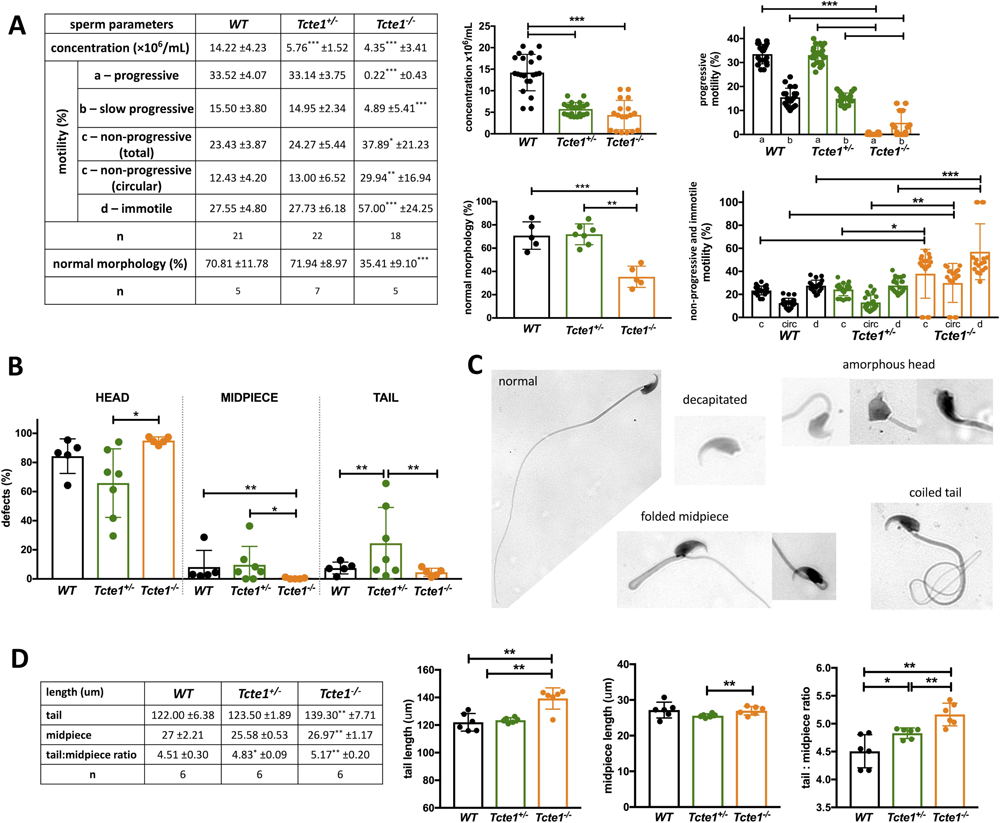
Characteristics of spermatozoa of *Tcte1* wild type (WT), hetero- ^(+/-)^ and homozygous mice ^(-/-)^. **A** Sperm parameters: concentration, motility and morphology comparison; statistical marks indicated differences according to WT animals; n – number of mice; **B** Frequency of morphological defects according to three part of the spermatozoa; **C** Examples of main sperm morphological abnormalities found (all abnormalities observed available in Supplementary file 7); Leica DM5500 microscope, objective 63x, CytoVision software; **D** Comparison of the mean length of: sperm tail, midpiece and ratio tail:midpiece; n – no. of animals, each with at least 100 spermatozoa analyzed; *** p < 0.0001, ** 0.0001 < p < 0.01, * 0.01 < p < 0.05

Increased mean length of tail and tail:midpiece ratio in *Tcte1^-/-^* animals (139.30 ± 7.71 um; 5.16, respectively) was measured when comparing to WT (122.00 ± 6.375 um; 4.51; p<0.002) and *Tcte1^+/-^* (123.50 ± 1.89; 4.83; p<0.008) (Figure 6D). Also, in the *Tcte1^+/-^* the midpiece length differed from *Tcte1^-/-^* animals (p=0.0334), followed by tail:midpiece ratio differences when compared to WT (p=0.0435).

### Sequencing of RNA (RNAseq) and STRING pathway analysis

RNA sequencing analysis of gene expression in testes of KO mice revealed statistically significant changes (p<0.05) in 21 genes, including 5 genes both for *Tcte1^-/-^* vs. *Tcte1^+/-^* and WT, 8 genes only for *Tcte1^-/-^* vs. WT, 7 genes only for *Tcte1^-/-^*vs. *Tcte1^+/-^*, and 1 gene both for *Tcte1^+/-^* vs. *Tcte1^-/-^* and WT (Figure 7). In *Tcte1^-/-^* animals vs. both: *Tcte1^+/-^* and WT, the expression level of 4 genes from kallikrein subfamily was decreased 2-8-fold, including genes involved in semen liquefaction and degradation of extracellular matrix proteins in the interstitial area surrounding Leydig cells of the adult mouse testes (*Klk1b27*, *Klk1b21*, *Klk1b24*), or neuronal activity (*Klk1b22*) (Figure 7; Supplementary file 12) [Michaelis *et al.,* 2017; Marques *et al.,* 2016; Du Plessis *et al.,* 2013; Fink *et al.,* 2007]. 2-fold decrease of expression of genes related to mitochondrial cytochrome oxidase subunits and asthenozoospermia was documented (*mt-Co2*, *mt-Co3*) (Figure 7) [Heidari *et al.,* 2016; Mostafa *et al.,* 2016; Baklouti-Gargouri *et al.,* 2013]. In a scheme: *Tcte1^-/-^*vs. WT, 8 genes revealed changes in expression level, including 4.7-fold rise for *Fetub* gene, which is known as required for egg fertilization (via specific inhibitor activity of the *zona pellucida* (ZP) ovastacin, the cysteine protease responsible for ZP2 cleveage) [Binsila *et al.,* 2021; Karmilin *et al.,* 2019], and plays also a role of a secreted factor during reprogramming [Bansho *et al.,* 2017].

**Figure 7:**
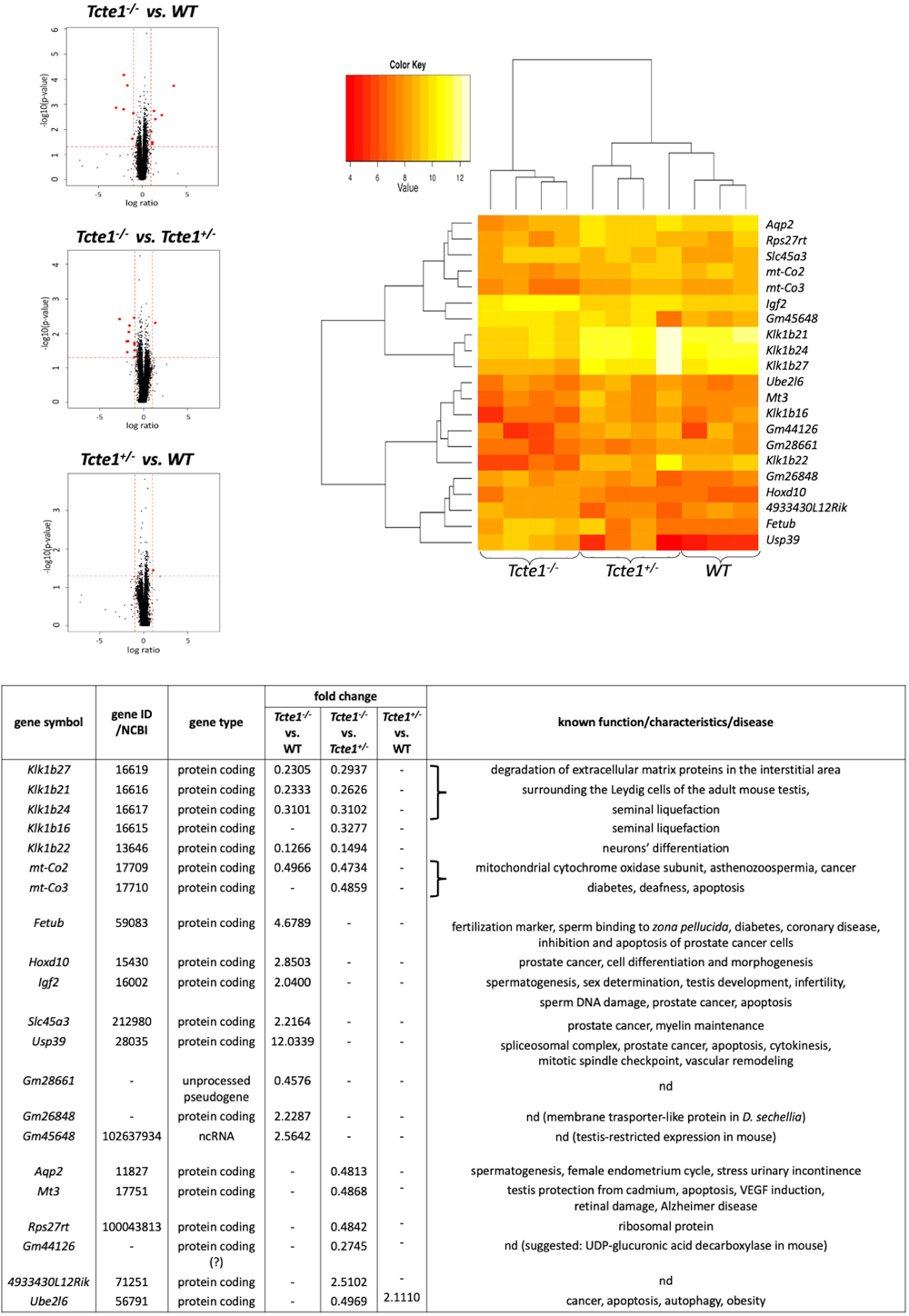
RNAseq results of changes in gene expression level found in testes of KO *Tcte1^+/-^* and *Tcte1^-/-^* mice vs. control (WT). Volcano plots: black dots indicated all analyzed genes within the genome, red dots represented genes with significantly changed expression level (followed by the heatmap and the table). Filtering criteria: p val <0.05, log ratio >1 or <-1 (adequate to fold change value of 2 (expression level increased) or 0.5 (expression level decreased)), expression level >8. nd – no data concerning function or disease.

Two-fold rise for *Igf2* gene (strongly linked to spermatogenesis, sex determination, testis development, infertility and prostate cancer; [Matsuzaki *et al.,* 2020; Cannarella *et al.,* 2020; Neirijnck *et al.,* 2019; Damaschke *et al.,* 2017]) has been also noticed. Igf2 and Klk proteins are enclosed in a common network consisted of proteins directly linked to fertility (Figure 8; Supplementary file 12; Supplementary file 13). One of them is Ambp, a fusion protein, which proteolysis generates bikunin – a protein which deficiency results in infertility, due to the diminished formation of the stable cumulus-oocyte complex required for maturation and ovulation [Sato and Kan,2001; Naz and Rajesh, 2005]. Bikunin is one of the elements of ITIH domains (Inter-α-Trypsin Inhibitor Heavy-chain), responsible for hyaluronian metabolism [Hull *et al.,* 2012; Salustri *et al.,* 2019]. ITIH is also linked to Plg, which has the protease activity and activates metalloproteins, such as: Mmp8 and Mmp9, also included in the network. Mmp8 is engaged in degradation/stabilization of occluding and blood-testis barrier [Chen *et al.,* 2018], while Mmp9 (with Igf1 and Igf2), is important for normal placentation [Ghosh *et al.,* 2011]. Igf1 and Igf2, are related to Igfbp3, which interacts with humanin expressed in Leydig cells. Humanin together with IGFBP3 and Bax prevents the activation of germ cell apoptosis [Jia *et al.,* 2015; Lue *et al.,* 2021]. What is important, expression of *IGFBP3* is regulated by *HOXD10* gene [Xue *et al.,* 2013; Pan *et al*., 2021], of which expression has been found with 2.9-fold rise in our study. *Hoxd10* is involved in prostate cancer processing, cell differentiantion and morphogenesis [Jonkers *et al.,* 2020; Mo *et al.,* 2017; Zhang *et al.,* 2019].

**Figure 8:**
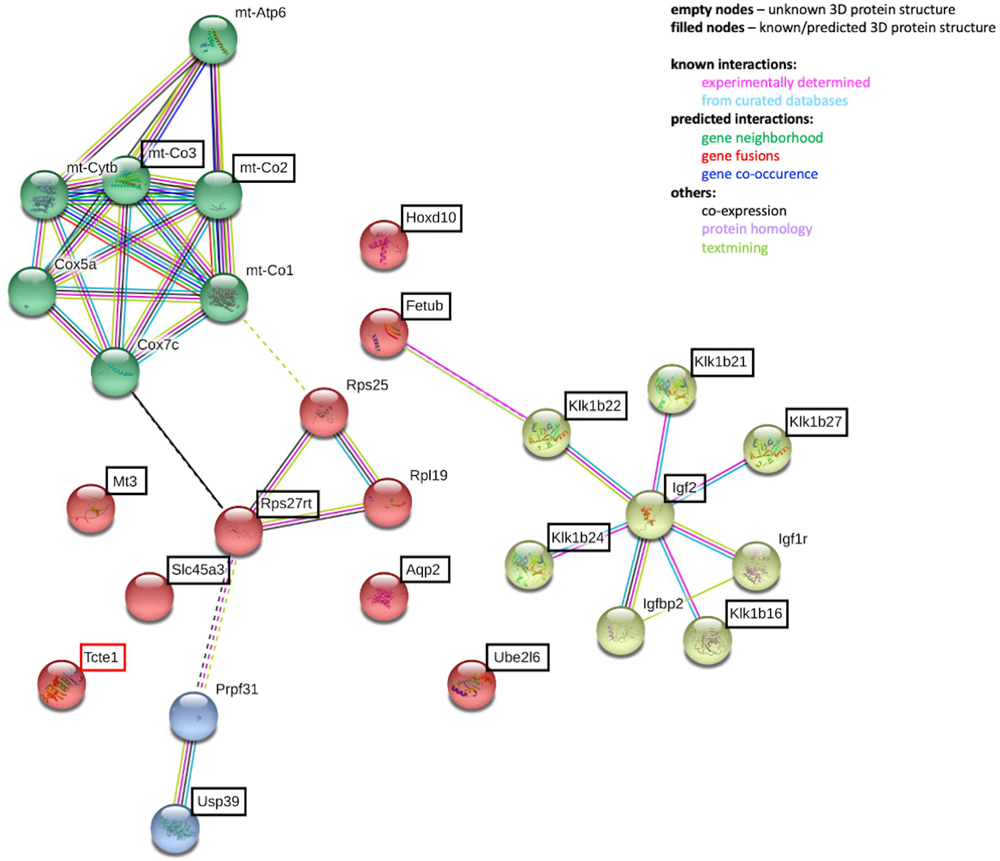
STRING analyzes of potential interactions between proteins with observed differences in gene expression level in RNAseq for mouse testis of the KO model for *Tcte1* gene. Genes revealed in RNAseq were bracketed. (STRING database, 24th Feb 2021).

Another gene with 2.2-fold expression rise is *Slc45a3,* a well-known marker of prostate cancer progression, when a loss of the protein determined poorer prognosis for cancer patients [Hernández-Llodrà *et al*. 2017; Pin *et al.,* 2017; Perner *et al.,* 2013]. An overexpression of another gene identified in our study (12-fold rise for *Usp39* gene) is linked with promotion of prostate or ovarian tumorigenesis. *Usp39* is also involved in pre-mRNA splicing, cytokinesis and spindle checkpoint working, and is crucial for apoptosis [Huang *et al.,* 2016; Yuan *et al.,* 2021; Zhao *et al.,* 2016].

Also, 2.2-2.5-fold rise for ncRNA genes (*Gm26848*, *Gm45648*; no data concerning their role) and 2-fold reduction for one pseudogene (*Gm28661*; also no data) were found (Figure 7). Following the latest data (November 2021) from Gene NCBI database, the Gm45648 has been annotated as one with the testis-restricted expression in mouse but no function has been suggested. The second ncRNA *Gm26848* found is suggested as protein coding organic cation transporter-like protein across membranes in *Drosophila sechellia*. There were also two other genes with expression change (3.6-fold decrease – pseudogene *Gm44126*, and 2.5-fold increase – *4933430L12Rik* probably involved in antisense transcription). ‘Gene’ database data showed that the *Gm44126* in mice is rather a protein coding gene with the function of UDP-glucuronic acid decarboxylase, and contains two domains involved in NAD(P)H processing. Thus, it can be suggested that this gene is as a novel candidate involved in mitochondrial respiratory chain. Our data with 3.6-fold expression decrease of this gene seem to fit into observed diminished functioning of mitochondrial machinery in *Tcte1^-/-^* mice.

Another three genes manifested 2-fold decrease of their testis expression when comparing *Tcte1^-/-^* vs. *Tcte1^+/-^*(but not to WT): *Aqp2* (aquaporin-2 involved in osmotic transportation of the water), with well-known role in spermatogenesis, female endometrium cycle and urinary incontinence; [Klein *et al.,* 2013; Ramli *et al.,* 2019], *Mt3* (metallothionein-3, committed in homeostasis and detoxification of toxic metals, also in testis) important in apoptosis regulation of Bax and Caspase pathways [Gao *et al.,* 2021; Honda *et al.,* 2010]), and *Rps27rt* (ribosomal protein) (Figure 5; Supplementary file 12). It was also found that Rps27rt may interact indirectly with Usp39 and mitochondrial respiratory complex linking the mitochondrial energy processing with apoptosis (Supplementary file 12). Last gene – *Ube2l6*, was found to have 2-fold increased expression level in testis in *Tcte1^+/-^* animals vs. *Tcte1^-/-^* and WT. Its role has been established as ubiquitin enzyme engaged in: stress damage response, cell cycle progression, embryo development, immune response, and factors linked to ATP machinery (Figure 7) [Huang *et al.,* 2016; Yuan *et al.,* 2021; Zhao *et al.,* 2016; Sandy *et al.,* 2020; Cai *et al.,* 2021]. Increased expression of *Ube2l6* was linked to degradation/suppression of cancer cells and affected downstream apoptotic factors (i.e., Caspase 3, Caspase 9, Bcl-2, Bax) [Li *et al.,* 2018].

### Immunofluorescence on sperm

An example of immunofluorescence *in situ* staining on mouse spermatozoa used to detect the N-DRC proteins has been presented in Figure 9. WT mouse spermatozoa showed Tcte1 protein presence in the sperm head nucleus, midpiece, and a part of the tail (Figure 9A). Immunofluorescent signal of Tcte1 in *Tcte1^+/-^*mice was similar to WT animals, while in *Tcte1^-/-^* was localized only in the nucleus (Figure 9A). It seems to confirm that in *Tcte1^-/-^* males the protein has not been produced properly, revealing only residual amounts in sperm head nucleus, and not transported to sperm flagella. The other N-DRC components (Drc7, Fbxl13 and Eps8l1) were localized in the sperm nucleus and among the whole sperm tail (Fbxl13, Eps8l1) or only within its fragment (Drc7), with exception of the midpiece (Figure 9B). Results obtained seem to suggest co-localization of evaluated N-DRC proteins in various combinations, not common among the midpiece and the terminal part of the sperm tail.

**Figure 9:**
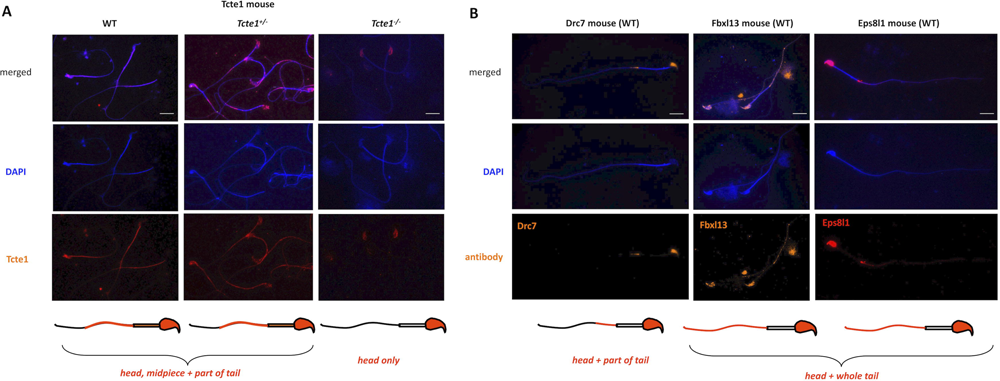
Immunolocalization of the nexin-dynein regulatory complex (N-DRC) proteins: Tcte1 (Drc5), Drc7, Fbxl13 (Drc6), and Eps8l1 (Drc3) within mouse sperm cells. **A.** Localization of Tcte1 in spermatozoa from wild type (WT), hetero- (^+/-^) and homozygous (^-/-^) animals; **B.** Localization of other proteins (Drc7, Fbxl13, Eps8l1) building the nexin-dynein regulatory complex (N-DRC), responsible for coordination of the dynein arm activity and stabilization of the doublet microtubules attachment; wild type (WT) spermatozoa. Antibodies: primary (all anti-rabbit 1:100; Biorbyt, UK): Tcte1 (orb357083); Drc7 (orb58695); Fbxl13 (orb678278); Eps8l1 (orb382538); secondary: goat anti-rabbit-AF594 conjugated, 1:500, ab150160, Abcam, UK. Fluorescent microscope: Leica DM5500, filters – DAPI, TxR, FITC, BGR, magnification 630 × (with immersion); software: LASX or CytoVision. Bar represents 10 μm.

### Sperm ATP level

Measurements performed in two time points (0 and +60 min.) revealed that in *Tcte1^-/-^* samples the luminescence level was approximately 2.4-fold lower when compared to WT and *Tcte1^+/-^* mice (p<0.0001) (Figure 10; Supplementary file 14). There were no differences between WT and *Tcte1^+/-^* males. Additionally, the decrease of the luminescent signal after 60 min. was similar in all three groups studied – approximately by 35% (WT: 35.27%, *Tcte1^+/-^*: 33.88%, *Tcte1^-/-^* 37.76%).

**Figure 10:**
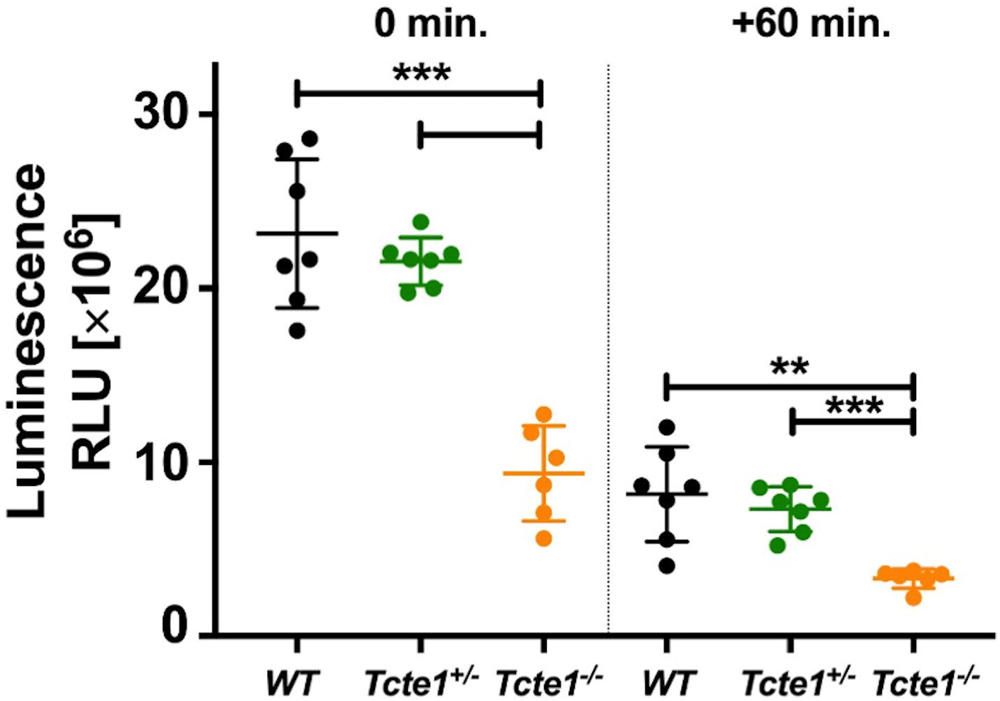
ATP level in wild type (WT), heterozygous (*^+/-^)* and homozygous (*^-/-^) Tcte1* mice. Measurement done in two time points (0 and +60 min.) showed decrease of approximately 35% of luminescence in all three groups. Mean measured values in homozygous group were approximately 2.4-fold lower. Dots – particular animals; long dash – mean value for the group; short dashes – standard deviation; RLU – relative luminescence unit. *** p<0.0001, ** 0.0001 < p < 0.01.

### Apoptosis in testicular tissue

Evaluation of the immunofluorescent staining results revealed Casp3-positive signals (indicating apoptosis) in spermatogonia only, in all three mouse genotypes (Figure 11A). In each zygosity the numbers of tubules and positively stained spermatogonia (intensity of the signal) were similar suggesting equal levels of apoptosis. Also lack of differences between zygosities were found in case of TUNEL assay, for all testicular cell types (Figure 11B). Apoptosis-positive signals were observed in all tubules for both techniques.

**Figure 11:**
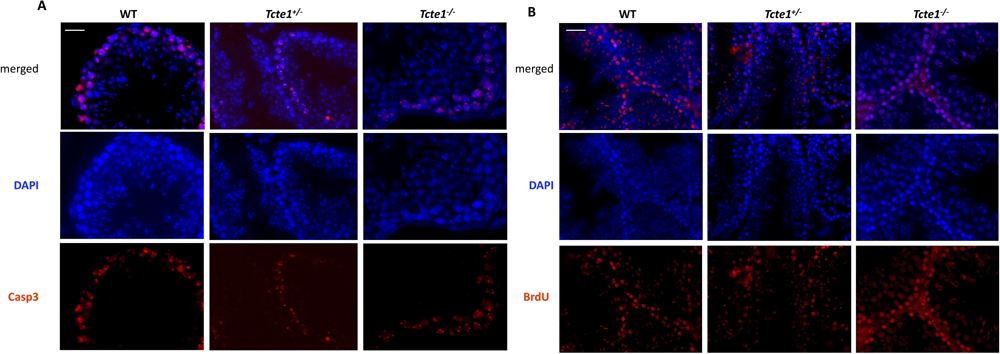
Immunolocalization of caspase 3 within mouse testicular tissue of KO *Tcte1* model. Caspase 3 is one of the apoptotic indicators becuase of its engagement in the induction, transduction, and amplification of intracellular apoptotic signals. Caspase 3 signals were found only on spermatogonia in all three genotypes evaluated (wild type (WT), hetero- (^+/-^) and homozygous (^-/-^)). No differences between the level of fluorescent signals were found between evaluated mouse genotypes, indicating similar level of apoptosis. Antibodies: primary (1:100) rabbit anti-Casp3 (Biorbyt, UK); secondary (1:500): goat anti-rabbit-AF594 conjugated (ab150160, Abcam, UK). Fluorescent microscope: Leica DM5500, filters – DAPI, TxR, magnification 630 × (with immersion); software: LASX. Bar represents 5 μm. Analysed in 3 independent males per zygosity, on 15-20 tubules each.

### Sequencing – human male samples

Potentially associated/important 6 heterozygous SNVs for *TCTE1* gene has been found in 15 out of 248 (6.05%) infertile males with drastically decreased semen parameters, ranging from azoospermia to severe oligoasthenozoospermia (Table 1). 3 variants were identified with ultrarare allele frequency (MAF ≤0.0005 in the general population; gnomAD) in three males with NOA: c.862C>T (p.Arg288Ter) predicted as stop-gained mutation leading to non-mediated decay, and thus to protein loss of function with high probability; c.185G>A (p.Arg62His), or c.1048G>A, (p.Gly350Ser), defined as missense ones and probably damaging. Next two SNVs were found as the rare missense ones with MAF <0.005: c.397C>T (p.Arg133Cys) (one NOA case) and c.470A>C (p.Glu157Ala) (four NOA cases), and were defined as probably damaging. The last 1 SNVs (1 NOA, 1 crypto-, and 5 with severe oligoasthenozoospermia) was not found in databases, and thus specified as the novel one (c.374T>G; p.Ile125Arg) with missense prediction (Table 1; Supplementary file 15).

**Table 1.**
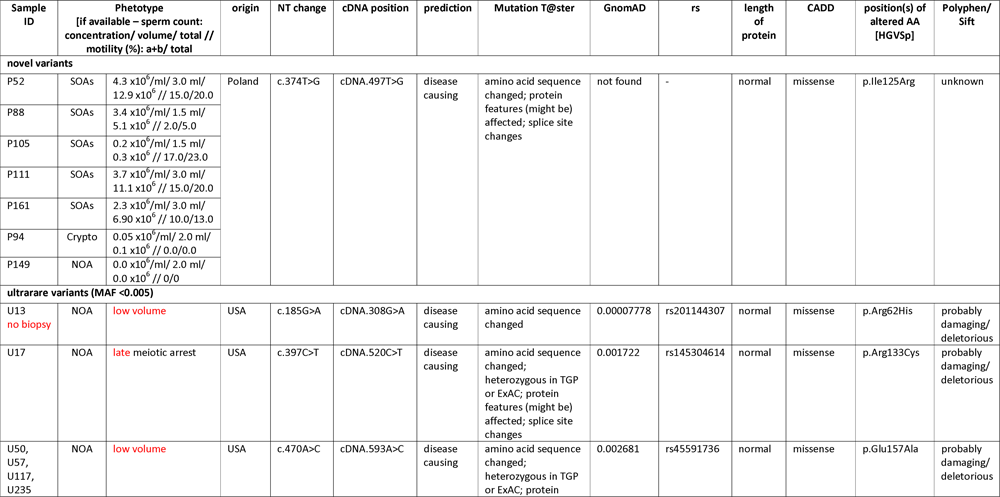
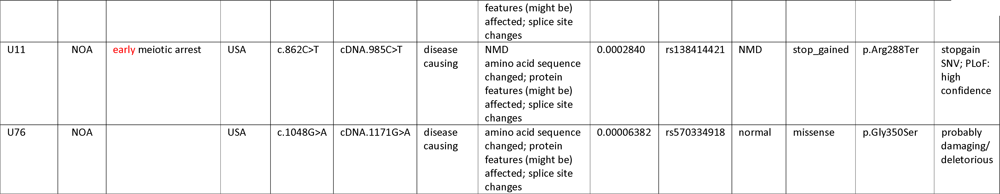
Potentially disease-causing novel and ultrarare heterozygous *TCTE1* variants found in patients with disturbed spermatogenesis. Results obtained from screening of n=248 participants. Minor allele frequency data obtained from GnomAD v2.1.1. SOAs – severe oligoasthenozoospermia; NOA – non obstructive azoospermia; Crypto – cryptozoospermia; NMD – non mediated decay; NT – nucleotide; AA – amino acid; SNV – single nucleotide variant; PLoF – Loss of Function probability; rs – Reference SNP accession number; CADD – Combined Annotation Dependent Depletion tool

### Protein prediction

To understand the molecular implications of identified mutations on the structure and function of TCTE1, we performed homology modeling experiments using the Phyre2, I-TASSER, and AlphaFold tools [Kelley *et al.,* 2015; Roy *et al.,* 2010; Yang *et al.,* 2015; Yang and Zhang 2015; Tunyasuvunakool *et al*., 2021]. TCTE1 is predicted to contain at least five tandem leucine-rich repeats (LRR) in its C-terminus [UniProt 2019], while no known three-dimensional protein folds have been assigned to its N-terminus. All homology searches yielded a polypeptide with tandem leucine-rich repeats with nearly 100% confidence (Figure 12A; Supplementary file 16). Modeling the N-terminal region of TCTE1 was more difficult; although both Phyre2 and I-TASSER confidently predicted a largely α-helical secondary structure for the N-terminal half of TCTE1 (Figure 12A; Supplementary file 16), neither tertiary structure was predicted with high confidence; indeed, comparison of the two demonstrates vastly different folds for the N-terminus from the programs. However, the confidence of the AlphaFold-generated model was high for the region just N-terminal to the LRR. Further, based on the predicted aligned error plot for calculated for its models, the confidence of the predicted structure of residues 100-200 was high relative to the position of residues 220-480, providing support for the modeled through-space three-dimensional interactions. That said, we interpreted these models very conservatively.

**Figure 12:**
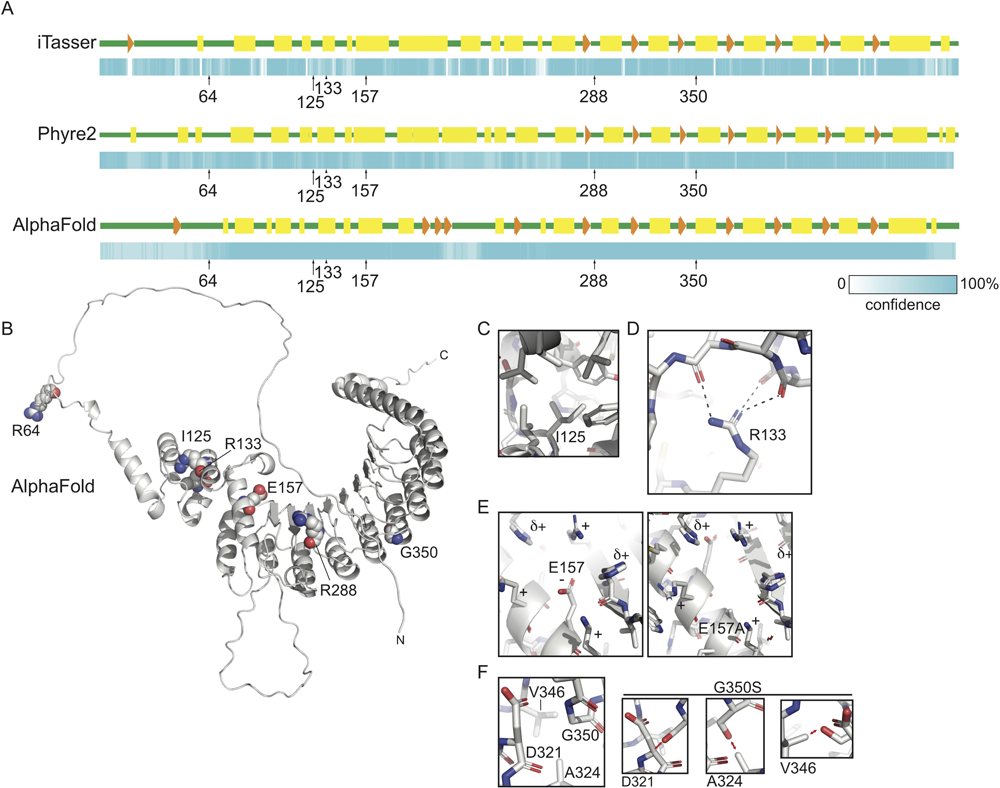
Homology modeling of TCTE1. **A** Secondary structure diagrams and associated confidence schema generated by I-TASSER [46,47,48], Phyre2[45], and AlphaFold [49,50] intensive mode. Positions of amino acids of interest are indicated. Green, predicted unstructured regions; yellow rectangles, predicted α-helices; orange arrows and arrowheads, predicted β−strands. Confidence scale ranges from white to cyan, with the probability of the secondary structure increasing with increasing intensity of cyan; **B** Cartoon diagram of homology model of human TCTE1 generated by AlphaFold with residues of interest shown as spheres and labeled. N, amino terminus; C, carboxy terminus; **C-F** Zoomed views of indicated residues and mutations: **C** Ile-125 resides deep within a hydrophobic pocket, which would be disturbed upon substitution with arginine; **D** Mutation R133C would impact the hydrogen bond interactions that organize the architecture of the globular domain N-terminal to the tandem leucine-rich repeats; **E** Mutation of Glu-157 to alanine would destabilize the charge of the surface of this globular region, likely leading to unfolding; **F** Gly-350 allows for the compact interactions of tandem α-helices of the LRR. Depending on the rotamer it adopted, a serine side chain in this position would sterically clash with residues D321, A324, or V346 in the neighboring helix.

We attempted to hypothesize how mutations of interest could affect the tandem LRRs and putative secondary structure of the N-terminus of the protein. Residue R64 resides in a predicted unstructured region and as such most likely is solvent exposed in the tertiary structure (Figure 12A, B). Mutation of this residue from arginine to histidine could therefore affect the surface potential of this disordered region, making the it less positively charged. Indeed, this mutation would be expected to impact the interactions that the N-terminus of TCTE1 can have with other molecules, including other members of the N-DRC. The interaction of microtubule-associated proteins, including the N-DRC, with binding partners is known to be largely charge-based [Kubo *et al.,* 2017; Drechsler *et al.,* 2019].

Three mutations of interest occur in the globular region just N-terminal to the LRR. The AlphaFold model clearly shows how mutation I125R would affect the structure of this region Figure 12A, C). Residue isoleucine 125 is predicted to be nestled within the hydrophobic core including L97 L109, L112, F91, W131, L197, Y130, and the methyl group of the T121 sidechain. A substitution of positively charged arginine for isoleucine in this position would not be accommodated in this hydrophobic region, likely resulting in local unfolding that could potentially propagate beyond the pocket. Likewise, residue R133 supports the structure of this putative globular domain by forming hydrogen bonds with the backbone of residues 109, 110, and/or 112, thereby possibly contributing to the overall organization of this domain and its positioning relative to the LRR (Figure 12D). Mutation of this residue to a cysteine would remove the hydrogen bonding groups supporting these interactions, likely leading to a local unfolding that could possibly propagate farther. Interestingly, residue E157 is predicted to interact with residues in the LRR, likely stabilizing positively charged residues around it, including several from the linker leading up to the first β-strand of the LRR (Figure 12B, E). Mutation of this residue to an alanine would lead to a tertiary destabilization, most likely affecting the overall structure of the N-terminal region of the LRR and the globular region just N-terminal to it.

Two mutations of interest affect the LRR helices. Mutation R288* results in premature truncation, eliminating most of the LRR region from the final protein product, although it is predicted that this premature termination codon would lead to nonsense mediated decay. Mutation G350S would be expected to result in the destabilization of inter-repeat interactions (Figure 12F), possibly even unfolding the protein from that position on; the lack of a side chain in glycine allows for the close approach of these tandem structural elements, whereas the addition of the serine side chain would abrogate the tight packing of residues, regardless of which rotamer it adopts. This could result in a restructuring its C-terminus and/or deforming the surface of the module, therefore affecting its interactions with its binding partners.

Since the N-DRC is conserved from single-celled eukaryotes to mammals, we took advantage of published cryo-electron tomography studies examining the N-DRC from model organism *Chlamydomonas* [Gui *et al.,* 2019]. This study was of great interest since it explicitly located the position of the C-terminus of DRC5 with a nanogold particle; importantly, it was consistent with earlier cryo-ET experiments positioning the N-DRC [Heuser *et al.,* 2009]. While the resolution of this structure is only 40Å, it was still useful for placing the modeled TCTE1 structure in the context of the entire N-DRC (Supplementary file 16B). Manual docking of the AlphaFold predicted TCTE1 structure is consistent with the cryo-EM tomography data [Gui *et al.,* 2019; Heuser *et al.,* 2009].

## DISCUSSION

In this study we have investigated the role of *Tcte1* in male infertility using mouse knockout model. Using homo-and heterozygous animals, their reproductive potential has been examined, noting no progeny by homozygous males. The size of the gonads varied between the zygosity of animals, followed by two semen phenotypes revealed within the sperm quality evaluation. RNAseq analysis of the mutant testicular tissue demonstrated the influence of *Tcte1* mutations on the expression pattern of genes related to mitochondrial cytochrome oxidase activity, apoptosis and spermatogenesis. The ATP level estimate of spermatozoa showed its decrease in homozygous males. Additionally, protein prediction modeling of identified variants in human samples revealed changes in the protein surface charge potential, leading to disruption in helix flexibility or dynamics, and thus, suggesting the disrupted TCTE1 interaction with its binding partners within the axoneme. The major findings concerning observed abnormal sperm tail beating in KO *Tcte1* mice were summarized in Figure 13. We have also identified and discussed novel functions of genes with testicular expression significantly disturbed in the *Tcte1* knockout. Therefore, this study contains novel and the most complete data concerning *TCTE1* role in male infertility, so far.

**Figure 13:**
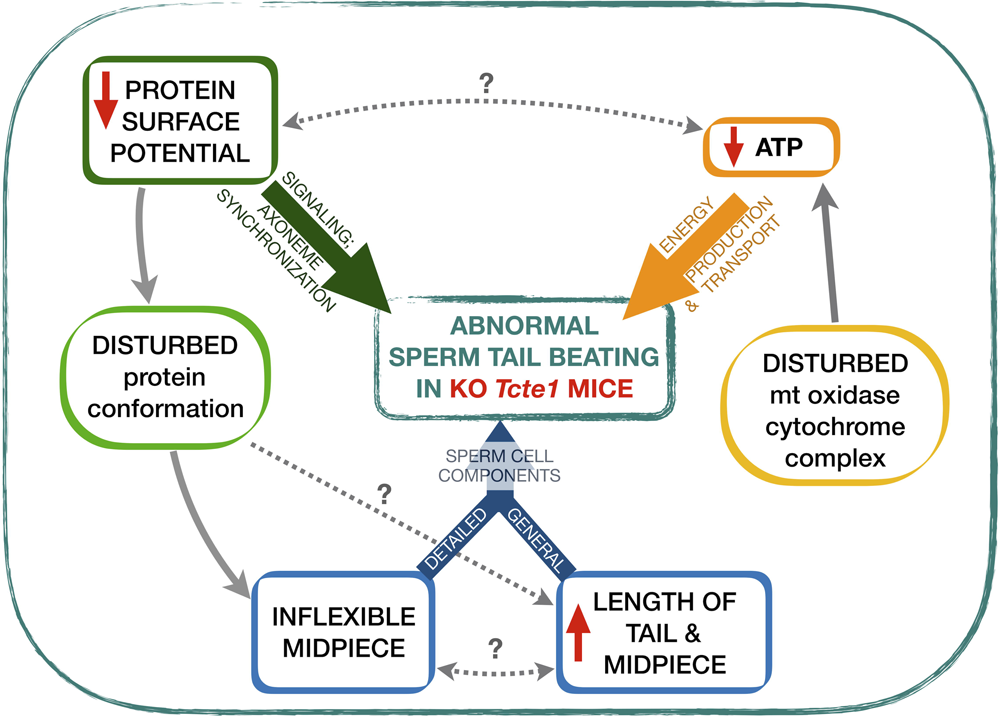
Summary of findings revealed for abnormal sperm tail beating in mouse knockout model of *Tcte1* gene, a component of the N-DRC complex in sperm tail. Three major groups of factors required for proper sperm tail beating were marked with bold colour arrows (green, yellow and blue). Documented connections (solid grey arrows) revealed on the basis of: RNAseq data for testis tissues, protein *in silico* predictioning for mutated Tcte1 protein, and measurements of ATP level and sperm cell components in mouse spermatozoa. Unresolved possible linkages (cause/effect) marked with question (?) mark.

### Phenotypes of homozygotes and heterozygotes

There was only one published study so far, concerning knockout mouse model of *Tcte1* [Castaneda e*t al.,* 2017]. It was shown that the expression of *Tcte1* starts at the stage of early haploid round spermatids and continues into the elongated spermatids. Authors have shown the infertility of homozygous males due to asthenozoospermia derived from limited and circular flagellar beating, followed by fertile heterozygotes. The study did not find any differences between homozygous and WT males in the testicular histology, followed by similar testis weight, sperm concentration and sperm morphology [Castaneda *et al.,* 2017]. We also have not found any structural differences in the testicular and epididymal tissues histology. However, the weight of the testes decreased significantly (∼41% in *Tcte1^-/-^,* 7% in *Tcte1^+/-^*), followed also by testis size reduction (by 22%) and lowered germ cell layer thickness in *Tcte1^-/-^* animals (Figure 4B, C). When comparing semen parameters (Figure 6A-C), we have found enormous decline in sperm concentration in both: *Tcte1^-/-^* (3.3-fold) or *Tcte1^+/-^* (2.5-fold) males, and double increase in abnormal sperm morphology in *Tcte1^-/-^* animals. Similarly, to Castaneda *et al*. [2017], we have documented the impaired sperm motility representing circular mode of the sperm tail beating. Also, the increased ratio of immotile spermatozoa in homozygous males was documented in our study. We can suggest that these discrepancies may result from various number of animals used for evaluation: 3 males in study of Castaneda *et al.,* [2017] vs. at least 18 per each zygosity in our study, when comparing semen parameters and tissue characteristics (except of sperm morphology evaluation with at least 5 animals used). Probably, also the differences in knockout construction used could be causative for observed variances: reporter-tagged insertion with conditional potential vs. the exon 3 deletion. Thus, we have observed two various phenotypes dependent on the zygosity of the animals: homozygous males manifested oligoasthenoteratozoospermia (OAT) and were infertile, while heterozygous males demonstrated oligozoospermia (O) and remained fertile. Data collected in International Mouse Phenotypic Consortium (IMPC; a repository of all available mouse phenotypes created, followed with the possible linkage to human disease defects; https://www.mousephenotype.org), concern only one *Tcte1* mouse study [Castaneda *et al.,* 2017], while we reveal an unmatched phenotypes of decreased sperm count.

We suggest that similar effect – decreased sperm count linked to *TCTE1* mutations, could be observed in humans. Our pilot mutation screening of 248 human infertile males with severely decreased sperm count (from oligo-, via crypto-, to azoospermia) revealed 6 heterogeneous SNVs (1 novel, 3 ultrarare, and 2 rare; all predicted as disease causing) with the total frequency of 6.05% (Table 1). We propose that molecular epidemiology studies of infertile patients should reveal the existence of homozygous or a compound heterozygous *TCTE1* variants with clear direct downgrading effect on spermatogenesis. However, we claim that also heterozygous variants can be causative, similarly to *SPINK2* which deficiency has been documented as responsible for oligoasthenozoospermia in heterozygous mice vs. azoospermia in homozygous animals, followed by the azoospermia in human homozygous brothers vs. their heterozygous father with oligozoospermia [Kherraf *et al.,* 2017]. It can be explained by the probable haploinsufficiency occurrence, when the single copy of the WT allele in heterozygous combination of which a variant allele is insufficient to produce the WT phenotype. Our observations, followed by the *SPINK2* data and data related to infertility (i.e.: P1/P2 protamines, *Pde8b*, *Trim28*, *Tcfl5*), and to the other diseases (i.e.: *PIKFYVE* in cataract, *NFIA*, *SCN1A* in neurological or epilepsy disturbances, *KLF1* in hemoglobin persistenece, *NR5A2* in pancreatic cancer), seem to confirm that the severity of the phenotype depends on the gene expression level [Jodar and Oliva, 2014; Mei *et al.,* 2022; Ogura *et al.,* 2022; Heshusius *et al.,* 2022; Valassina *et al.,* 2022; Sandhu *et al.,* 2021; Leal *et al.,* 2021; Tan *et al.,* 2020; Xu *et al.,* 2022]. We have documented variable phenotypes for a novel *TCTE1* variant c.374T>G (p.Ile125Arg) that has been documented in seven males, including: azoospermia, crypto-, and severe oligoasthenozoospermics. Thus, it seems to suggest the reasonable candidacy of haploinsufficiency as the mechanism responsible for observed phenotypic spectra also in case of the *TCTE1* gene. There is an only one study available documenting the *TCTE1^-/-^* in human case, so far [Zhou *et al.,* 2022]. Among 130 asthenozoospermic patients, authors have found frameshift mutation c.396-397insTC (p.Arg133Serfs*33) in one of them that resulted in premature translational arrest of the forming peptide. Successful birth delivery was available only after *in vitro* fertilization (IVF). What is important, authors also checked IVF rate in homozygous mice, but without pregnancy success. The unresolved question is whether homozygous *TCTE1* mutation in human causes only asthenozoospermia or rather the semen quality phenotype is mixed, including also abnormal morphology, but this was not evaluated by the authors.

### Kallikreins, Aquaporin 2

Our pathway analysis revealed a network including two genes with changed expression level in testis: *Igf2* (2-fold increased) and *Klk1b22* (8-fold decreased), supported by an interesting set of proteins linked to apoptosis and cellular processing (Figure 7; Figure 8; Supplementary file 12; Supplementary file 13). We can suggest that observed *Igf2* overexpression in our study can be one of the associated factors leading to decreased sperm count and testis parameters in evaluated KO mice model because of its known relation to spermatogenesis and testis development [Matsuzaki *et al.,* 2020; Cannarella *et al.,* 2020; Neirijnck *et al.,* 2019; Damaschke *et al.,* 2017]. Documented reductions can be supported by the observed constrained expression of *Klk1b22* – kallikrein mostly known from its neuronal activity; however, it cannot be excluded that we have revealed additional role of this protein, similar to the other members of the kallikrein family – role in semen liquefaction and degradation of extracellular matrix proteins in the interstitial area surrounding Leydig cells of the adult mouse testes (*Klk1b27*, *Klk1b21*, *Klk1b24*) [Michaelis et al. 2017; Marques *et al.,* 2016, Du Plessis *et al.,* 2013; Fink *et al.,* 2007]. Notably, *Klk1b22* expression is significantly enriched in testis. While expression of all four kallikreins was decreased 2.5-8-fold in our *Tcte1* KO model, indicating their participation in the reduction of sperm parameters measured (Fig.5).

We have also documented 2-fold decrease of expression of *Aqp2* (Aquaporin 2), involved in fluid resorption and water movement within the duct system, with known role in spermatogenesis [Klein *et al.,* 2013; Ramli *et al.,* 2019]. It was documented that in stallions Aqp2 is being expressed in Leydig cells, round and elongated spermatids [Klein *et al.,* 2013]. Also, this protein works within a network of molecules that are linked to cell membrane trafficking (i.e., Rab11a, Myo5b), followed by activity of protein kinases (Prkacb, Prkaca) that are known as regulators of cell proliferation, chromatin condensation/decondensation, and nuclear envelope disassembly/reassembly, and all is supported by the ATP metabolism (Hspa8) [Supplementary file 12] [Markou *et al.,* 2022; Wagner *et al.,* 2022]. Considering literature data, we suggest Aqp2 play an important role in reduction of the spermatids volume during spermatogenesis and in proper sperm morphology assessment and observed morphological differences between *Tcte1* mice zygosities in our study. However, such measurements require high-resolution microscopy, thus, we can only speculate Aqp2 role in our model.

### Disruptions in sperm cell structure

TCTE1 protein contains five leucine rich repeats (LRRs) that are known to be directly involved in receptor-mediated signaling on the structural basis for interaction between proteins [Ng *et al.,* 2011; Kobe and Kajava 2001]. It was documented that variants in genes encoding LRRs are commonly implicated in the pathogenesis of more than 60 human diseases [Ng *et al.,* 2011; Matsushima *et al.,* 2019]. LRR mutations mostly occur within regions that are protective for the hydrophobic core of the LRR domains. Thus, mutations in LRRs may affect the charge of the surface potential, leading to conformational changes of the protein or induction of protein cumulation. Our prediction model of potential changes in TCTE1 protein resulting from mutations found in human samples seems to point out the prospective impact into the interactions of the TCTE1 with other molecules (binding partners) known to be largely charge-base, including N-DRC elements or microtubule-associated proteins [Kubo *et al.,* 2017; Drechsler *et al.,* 2019]. In the case of TCTE1 protein observed amino acid substitutions promote the abnormal aggregation of TCTE1 protein, and decrease its structural stability, then probably leading to the observed disturbances. Beating motion of flagella is maintained by electrostatic cross-bridge between negatively charged tubulins and positively charged N-DRC [Kubo *et al.,* 2017]. Thus, changes in the charge potential of TCTE1 surface, which is one of the N-DRC components, seem to provide modified (reduced) flagellar beating, similarly as in case of mutations in other N-DRC element – DRC4 protein, of which changes in charge were essential for proper flagellar motility [Kubo *et al.,* 2017; Lewis *et al.,* 2016]. Also, the inflexibility of the midpiece observed in spermatozoa of *Tcte1* knockout mice seems to be a consequence of changed structure of LRRs in Tcte1 protein.

It is also known that the lengths of the sperm midpiece and tail may be crucial in determination of sperm swimming velocity, followed then by decreased fertilization rate [Firman and Simmons 2010; Gu *et al.,* 2019]. Available data showed that sperm with shorter midpiece may swim slower and were observed in asthenozoospermic cases [Firman and Simmons 2010; Gu *et al.,* 2019; Anderson *et al.,* 2002]. The meta-analysis of sperm shape features according to sperm competition theory has also shown, that in majority of vertebrates the longer sperm cell (including midpiece or flagellum) has been attributed to faster swimming, followed by the statement that the sperm size may be increased when sperm number is lowered [Lüpold *et al.,* 2020]. In our study we have documented longer sperm tails and midpieces in *Tcte1^-/-^* animals, that in fact were infertile. Thus, we can suggest that: (i) the observed longer sperm tails and midpieces may be co-responsible for aberrant motility observed, and (ii) the documented longer sperm tails and midpieces may be an indirect result of constrained sperm count observed, because of some sort of compensation for aberrant motility. However, this issue remains only a suggestion because of a restricted literature data relative to similar observations. Bennison et al. [2016] revealed than in a passerine bird the sperm tail velocity was correlated to total sperm length, but only to some point of the length value at which the velocity started to be decreased. On the other hand, Gu et al. [2019] have shown that in mammalian species there were some changes in the axonemal slopes when the flagellar length increases. The results obtained within our study, and supported by other research concerning sperm tail beating pathways, should be a good start point for creation of further mathematical computations for better understanding of the asthenozoospermia.

### Constrained energy metabolism

The flagellar motility is determined by coordinated activity of asymmetrically distributed dyneins in the axoneme. The driving factor for this activity is the hydrolysis of ATP released from OXPHOS system (Mitochondrial Oxidative Phosphorylation System) [Tourmente *et al.,* 2015; Ford 2006]. ATP is being produced in mitochondria placed within sperm midpiece. In our study ATP level measurements revealed its decreased level in sperm cells of *Tcte1^-/-^* mice, what can result in faulty dynein functioning, leading to asthenozoospermia via abnormal sperm tail beating. We have documented that *Tcte1* KO resulted in significant changes in expression level of testicular genes involved in mitochondrial energy transport. That leads to the conclusion that homozygous *Tcte1* mutation results not only in inflexible midpiece leading to asymmetric circular motility, as presented in this study and by others [Fung *et al.,* 2018], and then to asthenozoospermia, but also to aberrant functioning of mitochondrial OXPHOS machinery leading to disruption of ATP production. Thus, the sperm tail has also insufficient power for beating because of the decreased ATP content. Castaneda et al. suggested that the reasons of the observed ATP decrease may result from disturbances in a glycolytic pathway [Castaneda *et al.,* 2017]. It is already known, that glycolytic energy is enough for spermatozoa to survive [Nascimento *et al.,* 2008; Zhang *et al.,* 2022]. However, for sperm motility and then fertilization, the OXPHOS energy production is required [Tourmente *et al.,* 2015; Ford 2006; Zhang *et al.,* 2022]. In our study, RNAseq results clearly revealed the influence of *Tcte1^-/-^* mutation on genes involved directly in electron transport chain, such as: *mt-Co2* and *mt-Co3*, crucial components of the respiratory chain complex IV, one of the OXPHOS elements. Observed reduced expression seems to clearly evidence the decreased ATP production via disturbed electron chain transportation, and then revealing the sperm motility reduction observed in *Tcte1^-/-^* males, leading to observed asthenozoospermia. Detailed STRING analysis revealed definite interactions between both listed proteins identified in our study, and a list of other mitochondrial ones (mt-Co1, mt-Cytb, Cox5a, Cox6a, Cox7c, mt-Atp6; Supplementary file 12; Supplementary file 13) clearly related to proper processing of cytochrome energetic activity [Brischigliaro and Zeviani 2021; Ganetzky *et al.,* 2019]. Overexpression or mutations in *COX1*, *COX2*, and *COX3* were previously observed in infertile men with varicocele and OAT, or diabetes [Heidari *et al.,* 2016; Mostafa *et al.,* 2016; Perrotta *et al.,* 2012]. Additionally, it was shown that system of COX2 and prostaglandins plays key role in development of male gonad, spermatogenesis and steroidogenesis, especially in functioning of Sertoli and Leydig cells, and also was found to be significant in the background basis of idiopathic infertility status [Frungieri *et al.,* 2015]. It seems that in our study the reason of observed disturbed motility, represented as circular movement of spermatozoa, supported by increased ratio of immotile sperm cells in *Tcte^-/-^* mice, leading to asthenozoospermia, resulted from two factors: (i) inflexibility of sperm midpiece resulted from changed protein charge level in the Tcte1 protein, followed by elevated lengths of sperm tail and midpiece, and (ii) decreased amount of ATP produced, that may be an outcome of inability to get sufficient energy for proper sperm motility.

### Protection and/or compensation?

We have also documented an important role of the *Fetub* gene for the first time in the male testis. Besides of its known role in female fertility [Karmilin *et al.,* 2019; Bansho *et al.,* 2017], the latest study pay attention also in the context of prostate cancer is a detailed way [Zhan *et al.,* 2020]. It was found that overexpression of *FETUB* increases the apoptosis of prostate cancer cells, followed by the inhibition of their proliferation, migration and invasion, and also slower growth of tumor when compared to controls [Zhan *et al.,* 2020]. Thus, in the overexpression mode, the Fetub plays protective role for the prostate. It seems to explain results obtained in our study, where the *Fetub* expression was increased almost 5-fold in *Tcte1^-/-^* mice, but no tumor signs have been observed in the mouse testis at the tissue level. Zhan et al. [2020] have found the connection between *Fetub* overexpression, and induction of Bax (Bcl-2-associated X protein) and capases, resulting in promoting the apoptosis. It is known that the Bax/Bcl-2 induction/inhibition is directly related to the release of cytochrome c (Cyt c) in the caspase-dependent signaling [Dadsena *et al.,* 2021; Samaiya *et al.,* 2021; Li *et al.,* 2021]. And cytochrome c is one of the two electron carriers in electron transport chain responsible for transport directly to the respiratory chain complex IV [Yin and O’Neil 2021; Alvarez-Paggi *et al.,* 2017]. Due to Cyt c release, complex IV is essential for apoptosis and cell fate [Samaiya *et al.,* 2021; Li *et al.,* 2021]. Following of all of those facts, our results seem to reveal the connection between *Fetub* and diminished mitochondrial chain IV activity in mice testis, what have not been reported so far. It can be also supported by the fact of the known relation between FETUB and the inactivation of the PI3K/AKT signaling pathway (key player in regulation of cell proliferation and apoptosis in cancerogenesis), in which the Bax/Bcl-2 are involved [Zhan *et al.,* 2020].

The novel testis-protective role in our study can be also credited to three other genes with increased expression level in testicular tissue: *Hoxd10* (3-fold increase), *Slc45a3* (2.2-fold), and *Ube2l* (2-fold). Overexpression/activation of *HOXD10* has been already documented as inhibitor of tumorigenesis [Jonkers *et al.,* 2020; Mo *et al.,* 2017; Zhang *et al.,* 2019]. Expression of *Slc45a3*, known also as ‘prostein’, has been documented as weaker in cancer tissues [Hernández-Llodrà et al. 2017; Pin *et al.,* 2017; Perner *et al.,* 2013]. Elevated expression level of *Ube2l* is known to be connected with the degradation/suppression of tumor cells [Li *et al.,* 2018]. On the other hand, the 12-fold increased gene expression level for *Usp39* (ubiquitin specific peptidase 39) – an oncogenic splicing factor which expression is positively linked with cancerogenesis’ promotion in prostate or ovaries, while its silencing induces apoptosis and cell cycle arrest, has been noticed [Huang *et al.,* 2016; Yuan *et al.,* 2021; Zhao *et al.,* 2016].

Taken together data obtained for genes related to cancerogenesis and/or apoptosis described above, we can hypothesize that we observe an interesting novel network of interactions between two groups of genes that are reciprocally neutralizing potential influence onto the testis tissue. Mainly, in the first group we have genes with suggested novel anti-cancer role in the testis (*Fetub*, *Hoxd10*, *Slc45a3, Ube2l6*), which increased expression level inhibits the cancer cell development, supported by protective role of *Mt3*; while in the second corner of the ring the *Usp39* gene with elevated expression level (the most from all the observed changes) should lead to reveal some tumor-like changes within the testis tissue. Following the fact, that both: histological evaluation, as well as apoptosis stainings (Fig. 11), have shown no differences between hetero-and homozygous mutant mice, our hypothesis seems to be justifiable. On the other hand, there is a prediction that many of the genes (especially so-called cancer-testis associated antigens, CTA) related to spermatogenesis may function in tumorigenesis [Aurriere *et al.,* 2021; Yang *et al.,* 2020; Babatunde *et al.,* 2017; Wang *et al.,* 2016; Bode *et al.,* 2014; Cheng *et al.,* 2011; Nagirnaja *et al.,* 2018]. Due to known similarities between oncogenic and spermatogenic processes, especially in the energy metabolism, we can carefully suggest/hypothesize that in our KO model the genes with changed expression profile found, known as highly involved in tumorigenesis, may also be important in spermatogenesis. Obviously, there is a need for further studies in this direction to confirm these suggestions or to reveal the other (still unknown) mechanisms.

### Conclusions

We have documented that *TCTE1* gene is the next one that should be added to the ‘male infertility list’ of molecular background because of its crucial role in spermatogenesis and proper sperm functioning. The mutations in mouse *Tcte1* knockout model revealed two phenotypes, depending from zygosity with infertile oligoasthenoteratozoospermic homozygotes, and fertile oligozoospermic heterozygotic males, suggesting haploinsufficiency mechanism involvement. All collected data pointing that the Tcte1 protein is functioning in spermatogenesis and show how its mutations add the level of complexity to the clinical manifestation of decreased semen parameters. The disrupted energy machinery of the spermatozoa, followed by newly suggested roles of genes related to cancerogenesis and/or apoptosis (here in the context of their neutralizing/compensation effect on the testicular tissue), seem to underline the complex and devastating effect of the single-gene mutation, on the variety of testicular molecular networks, leading to reproductive failure. Further advanced studies are required to reveal the details of mechanisms responsible for the molecular pathways of the interactions between Tcte1 (the structural protein) and other non-structural networks of which expression have been changed remarkably in this study.

## Data Availability

All data generated or analyzed during this study are included in this published article and its supplemental information files. Supplementary files are deposited in Zenodo at https://doi.org/10.5281/zenodo.7330637
RNAseq data are available in GEO database (https://www.ncbi.nlm.nih.gov/geo/) under the accession number: GSE207805.
The results described in the publication are based on whole-genome or exome sequencing data which includes sensitive information in the form of patient specific germline variants. Information regarding such variants must not be shared publicly following the European Union legislation described in the following document: https://publications.jrc.ec.europa.eu/repository/bitstream/JRC113479/policy_report_-_review_of_eu_national_legislation_on_genomics_-_with_identifiers_1.pdf, therefore the access to raw data that support findings of this study are available from the corresponding author upon reasonable request.

https://doi.org/10.5281/zenodo.7330637

## ACKNOWLEDGEMENTS

We are grateful to Oliwia Kordyl, MSc, and Katarzyna Kurek, MSc (IHG PAS, Poznan) for their technical help in preparation of the samples for sequencing, genotyping, and immunofluorescence staining.

## Authors’ contributions

**MO** design of the study, manuscript drafting and editing, coordination of the tasks, data interpretation, gene selection, preparation of human blood and semen samples, mice: mating, genotyping, gross sections and preparation of biological samples, RNAseq analysis, immunofluorescence, Sanger sequencing, ATP measurements; **AM** gene selection, data interpretation, genotyping, RNAseq analysis, collection and preparation of human testis samples, WGS and Sanger sequencing, manuscript editing; **TS** bioinformatic analysis of RNAseq and WGS results; **NP** WES of azoospermia samples (incl. bioinformatics); **AB** protein prediction modeling and its description; **SB** preparation of: DNA for sequencing, RNA for RNAseq, screening of databases for RNAseq results; **MKa** collection of human blood and semen samples, semen analyses of human and mouse samples, detailed morphology of mouse spermatozoa; **JS-Z** and **HJ** histopathology of mouse tissue sections; **PJ** recruitment of patients and their medical history; **ANY** gene selection, interpretation of azoospermia WES results, data interpretation; **MKu** data interpretation, gene selection, recruitment of patients and their medical history, correction of the manuscript, funds collection and supervision. All authors critically reviewed and approved the final version of the manuscript.

## LARGE SCALE DATA

All data generated or analyzed during this study are included in this published article and its supplemental information files.

RNAseq data are available in GEO database (https://www.ncbi.nlm.nih.gov/geo/) under the accession number: GSE207805.

The results described in the publication are based on whole-genome or exome sequencing data which includes sensitive information in the form of patient specific germline variants. Information regarding such variants must not be shared publicly following the European Union legislation described in the following document: https://publications.jrc.ec.europa.eu/repository/bitstream/JRC113479/policy_report_-_review_of_eu_national_legislation_on_genomics_-_with_identifiers_1.pdf, therefore the access to raw data that support findings of this study are available from the corresponding author upon reasonable request.

## LIMITATIONS, REASONS FOR CAUTION

In the study, the *in vitro* fertilization ability of homozygous male mice was not checked.

## WIDER IMPLICATIONS OF THE FINDINGS

This study contains novel and the most complete data concerning *TCTE1* role in male infertility. *TCTE1* gene is the next one that should be added to the ‘male infertility list’ of molecular background because of its crucial role in spermatogenesis and proper sperm functioning.

## Supplementary files legend

**Supplementary File 1: Vectors used for knockout creation of Tcte1 gene.**

**Supplementary File 2: Real time PCR conditions used for determination of relative expression level of *Tcte1* gene in mouse knockout animals.**

**Supplementary File 3: Reproductive potential of KO *Tcte1* mice.** Mating combinations and their results including: time of breeding, number of litters and pups, number of male and female pups, and genotypes observed in pups of each combination.

**Supplementary File 4: Genotyping protocol for *Tcte1* knockout model.**

**Supplementary File 5: Characteristics of antibodies used for N-DRC complex staining in mouse sperm.** Human antigen sequence identity between human and mouse orthologs was also shown (blast comparison; information from Biorbyt producer).

**Supplementary File 6: Schematic models of exon 3 deleted in KO mice. A** Tcte1 protein exon 3 (deleted in KO) prediction of 5 top models (software: I-TASSER). C-score is the confidence score of the prediction. C-score ranges [0-1], where a higher score indicates a more reliable prediction; TM-score is a measure of global structural similarity between query and template protein., RMSD - root-mean-square deviation of the average distance between the atoms. **B** The deletion sequence used for modeling, including exon 3 (yellow).

**Supplementary File 7: Examples of HE staining on testicular tissue.**

**Supplementary File 8: An example of circular type movement pathway of spermatozoa from *Tcte1^-/-^* male mice.**

**Supplementary File 9: An example of sperm cells motility in *Tcte1^+/-^* male mice.**

**Supplementary File 10: An example of sperm cells motility in *WT* male mice.**

**Supplementary File 11:** Detailed parameters of mouse sperm morphology. Statistically significant values: p < 0.05; ns – statistically not significant (p ≥ 0.05). For samples with normal distribution (positive Shapiro-Wilk normality test), t-test with Welch’s correction was applied; for samples without normal distribution (midpiece and tail defects), Kolmogorov-Smirnoff test was applied (GraphPad Prism v. 7.0e). n=200 spermatozoa were analyzed per each mouse; for WT and *Tcte1^-/-^* n=5 animals were evaluated, while for *Tcte1^-/+^* the number of animals was n=7. Spermatozoa stained with the classic Papanicolau staining procedure. Bright field microscope: Leica DM5500, magnification 630x (with immersion), software: CytoVision.

**Supplementary File 12: STRING analyzis data for each gene with changed expression pattern in *Tcte1* KO mouse model.**

**Supplementary File 13: STRING analyzis of potential interactions between proteins with observed major differences in gene expression level in mouse testis of the homozygous *Tcte1* mice (Usp39, Fetub, Klk1b22).** According to genes revealed in RNAseq analysis (marked with arrows), two groups of interactions were formed for genes with highest expression changes (red arrows); (STRING database, 11th March 2021).

**Supplementary File 14: ATP measurement data obtained for KO Tcte1 male mice.** wt = wild type, het = heterozygous, ho - homozygous. RLU = relative luminescence unit.

**Supplementary File 15: Examples of potentially disease-causing rare heterozygous *TCTE1* variants found in patients with disturbed spermatogenesis.** Results obtained from screening of n=248 participants. **A** novel variant (Sanger sequencing); **B** ultra rare variants (WES).

**Supplementary File 16: The C-terminal region of TCTE1 was consistently modeled and fits well in published maps. A** Superposition of the cartoon representation of three models of human TCTE1 generated by Phyre2 [45] intensive (green) or fast mode (grey), I-TASSER [46,47,48] (yellow), and AlphaFold [49,50] (cyan). N-and C-termini are labeled. **B.** Manual docking of the model of full-length TCTE1 generated by AlphaFold in maps calculated from a cryo-electron tomography experiment performed on *C. reinhardtii* using nanogold to locate the C-terminus of DRC5 (EMD-20821) [23].

